# A *FLOT1* host regulatory allele is associated with a recently expanded *Mtb* clade in patients with tuberculosis

**DOI:** 10.1101/2022.02.07.22270622

**Authors:** Yang Luo, Chuan-Chin Huang, Qingyun Liu, Nicole Howard, Xinyi Li, Junhao Zhu, Tiffany Amariuta, Samira Asgari, Kazuyoshi Ishigaki, Roger Calderon, D. Branch Moody, Leonid Lecca, Sarah M. Fortune, Megan B. Murray, Soumya Raychaudhuri

## Abstract

The outcome of infectious diseases may depend on the interaction between human and pathogen genomic variations. We explore this relationship in tuberculosis (TB) by conducting a genome-to-genome (g2g) study of paired genomes from humans and the infectious agent *Mycobacterium tuberculosis* (*Mtb*) in 1,556 Peruvian TB patients. We identified a significant association between a human variant in the *FLOT1* gene and a unique *Mtb* Lineage 2 (L2) subclade. The host allele affects *FLOT1* expression in multiple tissue and cell types including lung, the primary site of TB disease. Phylogenetic analysis shows that the *Mtb* subclade has expanded rapidly in Peru since its emergence in the 1950s. Unbiased phenotypic profiling demonstrates that strains from the interacting *Mtb* subclade display different redox metabolism from other L2 strains. This study presents clear evidence that human and bacterial genetic variation interact together to produce different clinical outcomes.

---

Infectious diseases account for much of the burden of all diseases worldwide, but their host genetic architecture is less well understood than many other types of complex traits, despite having a comparable genetic heritability(*1, 2*). Infectious diseases occur only when pathogenic organisms infect a human host, and thus they are unique in that the effect of human risk alleles may be influenced by genetic variation within the pathogen. In TB, the pathogenic organism *Mtb* has co-existed with humanity for millennia, and currently colonizes one-fourth of the population worldwide(*3*). In 2019 alone, approximately 10 million patients were diagnosed with TB, and 1.4 million succumbed to it(*4*).

Human genome-wide association studies (GWAS) by us and others have identified only a few confirmed disease alleles in TB. Most TB risk alleles appear to be unique to populations with restricted geographical locations(*5*–*10*). One possibility is that human alleles may predispose individuals to clinical disease differently depending on the *Mtb* strain, which varies from place to place in the world. If true, this situation could appear as a statistical interaction between host alleles with *Mtb* alleles(*11, 12*). Early studies in TB have explored the possibility that human host alleles may have specific associations to subphenotypes, such as disease severity and age of onset, in a *Mtb* lineage specific manner(*13, 14*). A full unbiased variant-to-variant search may have the potential to reveal heterogeneity beyond established *Mtb* lineages and may implicate novel human risk loci. Scaling of sequencing and genotyping technologies has now made an unbiased examination possible. Here, we use a genome-to-genome (g2g) approach to examine both genomes comprehensively in an unbiased fashion to find genetic determinants of host and pathogen interactions in progression to TB pulmonary disease.

## Results

We hypothesized that host genetic variation can predispose certain individuals to a higher risk of disease from specific bacterial lineages or clades, including those that are not yet defined. If these differences in lineage frequencies across human hosts are genetically driven, we should be able to identify an association in TB patients between host and bacterial genetic alleles.

To this end, we collected both human genotype and *Mtb* whole-genome sequences (WGS) from 1,556 TB patients in Lima, Peru (**Fig. 1, Table S1**), where the TB incidence rate is one of the highest in South America(*15*). After quality control, we obtained 676,110 genotyped human variants with a minor allele frequency ≥ 1%. We focused our g2g analysis on 2,298 out of 45,831 called *Mtb* variants with an allele frequency between 5% and 95%, since statistical power for rare *Mtb* alleles would be limited.

**FIG 1.**
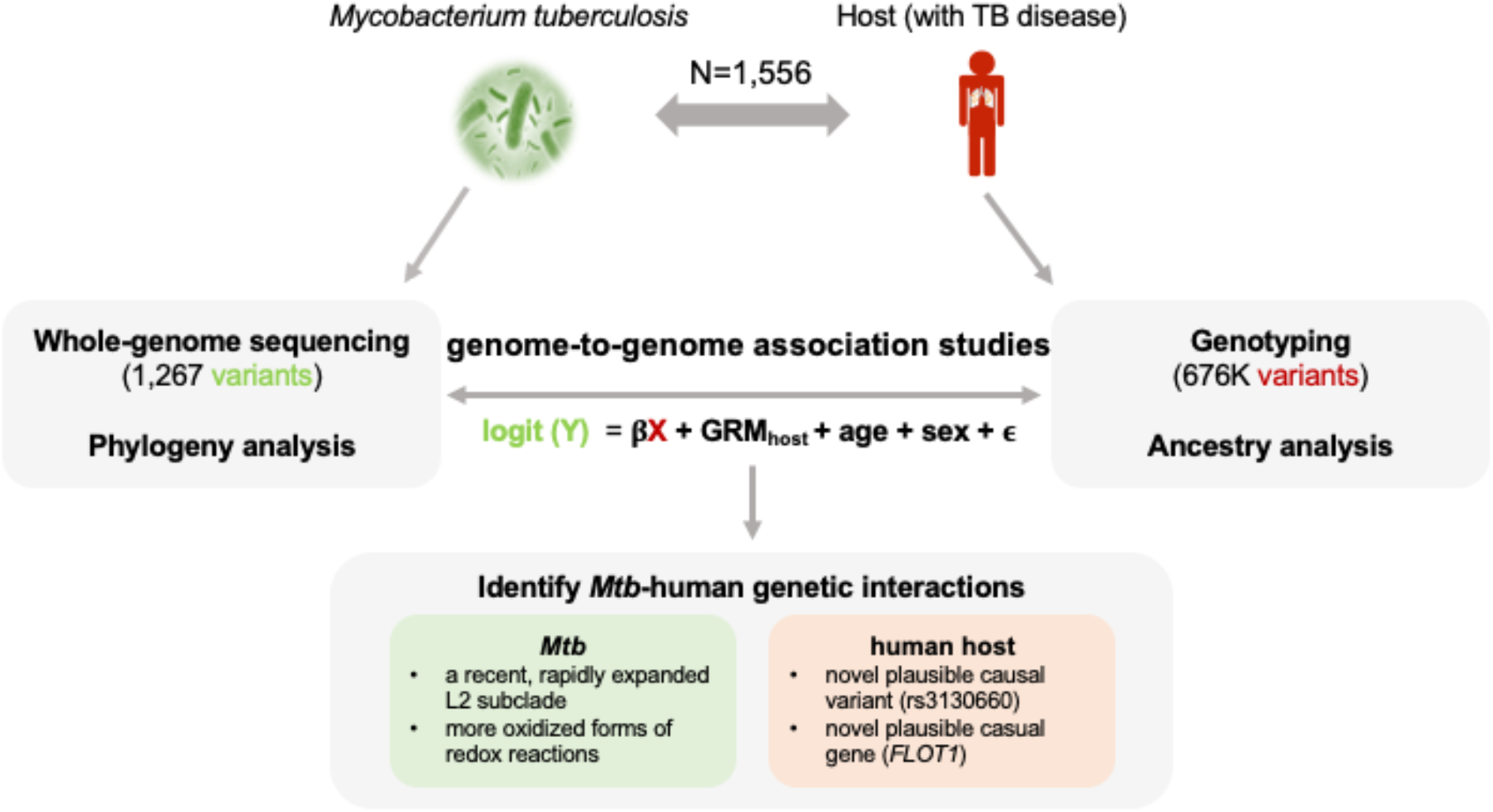
Study design schematic. We obtained DNA from 1,556 Peruvian individuals with TB disease and cultured pathogens to perform host genotyping and *Mtb* WGS. The genotype of each common *Mtb* variant was considered as the response variable (Y: 0 or 1), and the genotype of each host variant was the independent variable (X: 0, 1 or 2), resulting in one test per host SNP-*Mtb* SNP pair. This g2g approach identifies alleles that show evidence of host-pathogen interaction. We studied the identified genomic interaction by pinpointing plausible causal host g2g variants and genes, as well as characterizing g2g *Mtb* variants in the phylogenetic tree and experimental assays.

We tested whether the co-occurrence of each pair of common bacterial and human variants was higher than expected by performing a mixed effects logistic regression. We considered the presence or absence of each common *Mtb* SNP as a binary trait and assumed an additive model correcting for human cryptic relatedness, population structure, age and sex. To reduce the computational burden, we only included 1,267 (out of 2,298) common *Mtb* SNPs that were not in near perfect linkage (Pearson r^2^ < 0.99) in the association tests. Similar to performing GWAS on two correlated traits (e.g., blood pressure and cholesterol level), we allowed correlation in our outcome variables (*Mtb* SNPs) and did not correct for *Mtb* structure in our model. As a result, a single host variant can be associated with multiple *Mtb* variants. In total, we ran > 850 million regression models between every common *Mtb* and host variant pair (**Fig. 2A**). We examined genomic inflation factors (λ_gc_) of each of 1,267 GWAS, and observed no inflation of test statistics (median λ_gc_=1.01, **Fig. S1**), suggesting that our model is robust to false-positive findings. We used permutations to rigorously determine a significance testing threshold accounting for correlated tests and any inflation that might be present (P < 6.55 × 10^− 12^ assuming α = 0.05 (**Fig. S2**).

**FIG 2.**
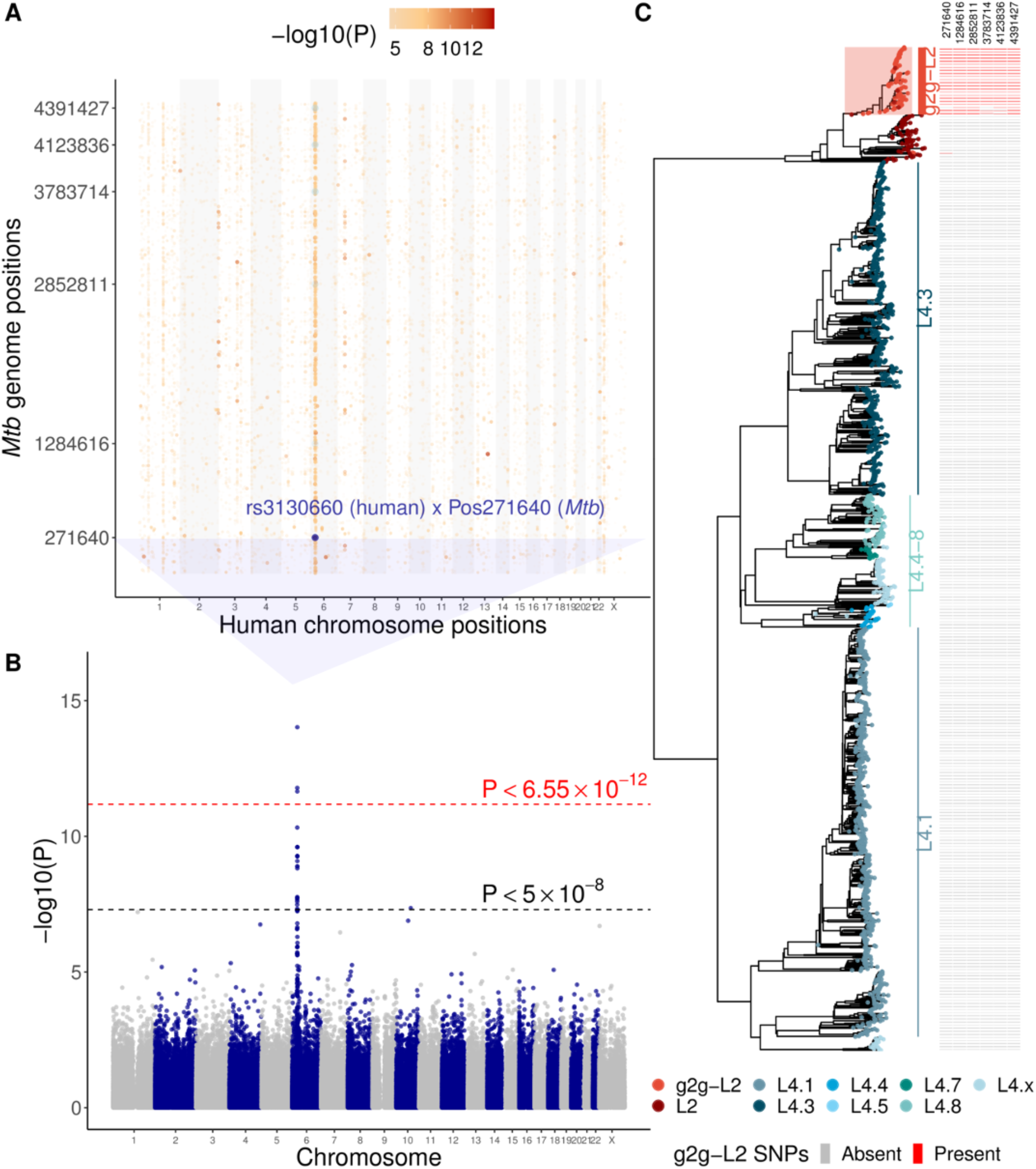
Human-to-*Mtb* genome-wide association study in 1,556 tuberculosis patients. (**A**) Grid plot summarizing the genome-to-genome analysis. The x-axis denotes position within the human genome with alternating colors for each chromosome. The y-axis denotes position within the *Mtb* genome. Point colors represent the association p-value (-log_10_(P)) from the logistic regression. The most significant host-*Mtb* pair association is indicated. Six randomly chosen *Mtb* variants in tight linkage (Pearson r^2^ > 0.8) with position 271640 are shown in light blue, indicating that the same human variant rs3130660 is significantly associated with multiple *Mtb* positions. (**B**) Manhattan plot of the GWAS analysis when treating genotypes of *Mtb* position 271640 as the outcome. The red dotted line denotes the genome-wide significance threshold (P < 6.55 × 10^−12^) derived from permutations. The black dotted line denotes the standard genome-wide significance threshold (P < 5 × 10^−8^). (**C**) A maximum likelihood phylogenetic tree inferred from 13,981 variants of 1,555 Peruvian *Mtb* isolates (excluding one Lineage 1 sample for visualization purposes). Branch colors represent the inferred lineages. Filled squares on the right indicate the presence (red) or absence (gray) of the six *Mtb* variants identified in the g2g analysis and highlighted in (**A**).

### Genome-to-genome study identifies human-Mtb genomic interactions

We identified an association between an intronic human variant, rs3130660, located in the *6p21* region on chromosome 6 and a phylogenetic marker (Position 271640) of a subclade of L2 *Mtb* (OR = 1.39, 95% CI: 1.33-1.44, P = 9.24 × 10^−15^, **Fig. 2B**). Individuals with each rs3130660 A allele were ∼40% more likely to be infected with *Mtb* strains carrying the interacting *Mtb* variant marker. We noted that the associated *Mtb* variant is in tight linkage with 78 of the 2,298 common *Mtb* variants (Pearson r^2^ > 0.8, **Table S2)**.

To better understand these 79 *Mtb* variants, we constructed a maximum likelihood phylogenetic tree using WGS of 1,556 collected isolates. In this analysis, we looked at 13,981 *Mtb* variants which included both the 2,298 common variants along with 11,683 variants that passed stringent quality control. We classified the strains into well-defined *Mtb* lineages, including L2 and L4.1-8 using previously defined lineage defining markers(*16*). We observed that previously defined lineages fell within distinct branches of the phylogenetic tree (**Fig. 2C**). Strikingly, we observed that all 79 *Mtb* SNPs were present in the same L2 subclade of the phylogenetic tree. We henceforth define the clade by the presence of the g2g *Mtb* variant (Position 271640) as the g2g-L2 clade.

### Human allele associated with Mtb diversity colocalizes with FLOT1 expression in lung and other tissue types

The strongest association in the human genome is an intronic variant (rs3130660) of the *FLOT1* gene (**Fig. 3A**). FLOT1 is a lipid raft-associated scaffolding protein that plays a role in membrane trafficking and phagosome maturation(*17, 18*). We investigated whether this SNP modulates the expression levels of nearby genes by performing an expression quantitative trait loci (eQTL) analysis. Using the Genotype Tissue Expression (GTEx release v8(*19*)) database in lung, we observed that this SNP was strongly associated with *FLOT1* expression (effect size = 0.28, P = 2.22 × 10^−16^, **Fig. 3B, Fig. S3**). Each rs3130660 A allele corresponds to an 1.32-fold increase in *FLOT1* expression in the lung. The most strongly associated variant with *FLOT1* expression in GTEx was rs3132610 which is in tight linkage with rs3130660 (r^2^ = 0.90 in 1KG European population(*20*), r^2^ = 0.77 in this Peruvian cohort). To understand whether the rs3130660 allele corresponds to the signal reported in GTEx, we applied a colocalization analysis using the *coloc* software(*21*). We note that there are differences in genetic ancestry between our study and GTEx, which are predominantly individuals of European ancestry, and this might reduce the power to detect colocalizing signals across datasets(*22*). We also note that there are multiple independent alleles influencing *FLOT1* expression(*19*), which might further reduce power to detect colocalization (**Fig. 3B**). Despite this, we observed strong evidence of colocalization in the lung (posterior probability = 0.98). Consistent with the colocalization analysis, when examining 14,976 SNPs overlapping with eQTL summary statistics of *FLOT1*, we observed strong correlation between association statistics of *FLOT1* expression in lung and infection with g2g-L2 strain (Pearson r = 0.89, **Fig. S4**). We then looked at other cell and tissue types and assessed colocalization signals between *FLOT1* and other 49 protein coding genes within the 700kb window of rs3130660 (**Fig. 3C, Table S3**). Using 108 RNA sequencing datasets included in the eQTL catalog release 4(*23*), we found evidence of colocalization between the identified interacting host SNP and *FLOT1* eQTL association in 15 cell and tissue types (posterior probability > 0.8), including T cells (posterior probability = 0.99) and adipose (posterior probability 0.96, **Fig. 3D, Fig. S5**). These results provided strong evidence for colocalization between the host variant associated with the g2g-L2 strain and host variants associated with *FLOT1* expression levels in multiple tissues and cell-types. This suggests that *FLOT1* may be the causal human gene interacting with the *Mtb* genome.

**FIG 3.**
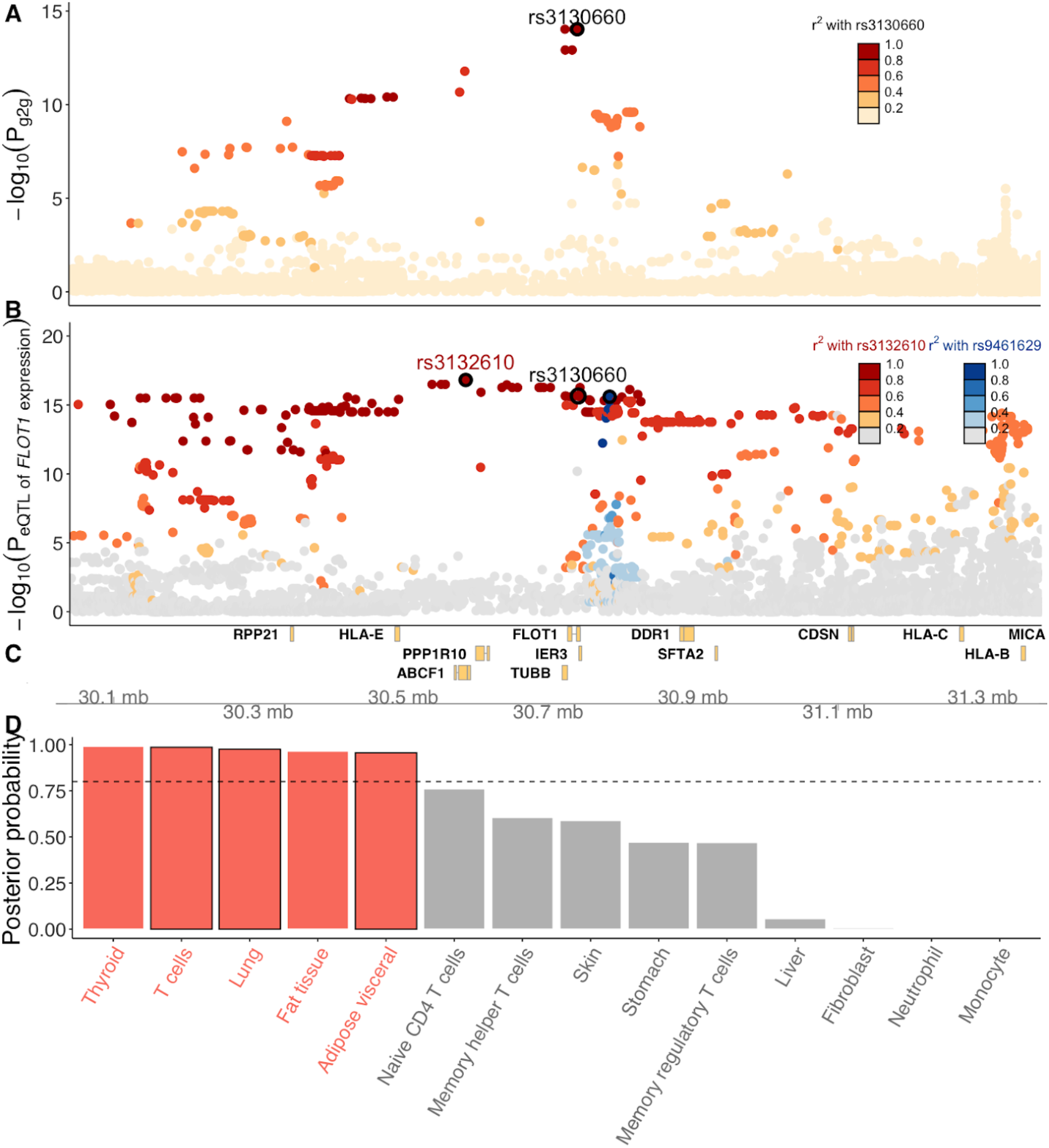
Association in the human host *FLOT1* locus. (**A**) Manhattan plot of the g2g association analysis in the *FLOT1* locus. The y-axis shows the -log_10_(p-values) obtained using a linear mixed model. Point colors represent the LD r^2^ with the lead host variant (rs3130660) in this Peruvian cohort. (**B**) Manhattan plot of the *FLOT1* eQTL. The x-axis shows the human genomic positions. The y-axis shows the -log_10_(p-values) for the eQTL association between host variants and *FLOT1* expression level in the lung (obtained in GTEx). Point colors represent the LD r^2^ between either of the two eQTL SNPs of *FLOT1* expression (rs3132610, circled in red and rs9461629 circled in blue) (**C**) Genomic locations of all genes in the *FLOT1* locus in GRCh38. (**D**) A barplot of all posterior probabilities of shared causal variants between the g2g and *FLOT1* eQTL signals in 13 cell and tissue types selected among 108 bulk RNA-sequencing datasets. Completed colocalization results in 108 datasets are provided in **Table S3**.

To test whether this result reflected social mixing patterns within the community or a true biological association, we repeated our analysis adjusting for year of TB diagnosis and *Mtb* population structure (as reflected by principal components), neither of which substantially affected our results (**Fig. S6**). Notably, the prevalence of the g2g-L2 clade remained consistent over time (**Fig. S7**) and over geographical space (**Fig. S8**), reinforcing our conclusion that the reported signals are independent from these covariates.

We considered the possibility that the observed association between a host allele and the *Mtb* g2g-L2 subclade was driven by host ancestry (**Fig. S9**) or *Mtb* lineage (**Fig. S10**). We examined host alleles for associations to the two common *Mtb* lineages in Peru (L2 and L4), and found the strongest association was between the same *FLOT1* allele and the L2 lineage, but that this association was substantially weaker than with the g2g-L2 lineage (rs3130660, OR = 1.37, 95% CI: 1.29-1.44, P = 7.04 × 10^−9^, **Fig. S11**). A conditional analysis including g2g-L2 obviated this association (P = 0.97). In contrast, adding L2 as a covariate to our original model did not obviate the association wit g2g-L2 (P = 8.15 × 10^−8^). This suggested to us that the observed association between rs3130660 and L2 was driven by the g2g-L2 subclade rather than L2 more broadly. We also tested the effect of five human host ancestry principal components on *Mtb* genomic variants (1,267 tests) and the association of host ancestry principal components on two *Mtb* lineages. We observed no significant associations, suggesting that host genetic ancestry is not associated with the variations in the *Mtb* genome in Peru.

### Mtb g2g-L2 subclade is recently expanded in Peru and transmits differently than other L2 strains

To interrogate the population dynamics of local *Mtb* L2 strains identified in the g2g analysis, we used 178 L2 isolates from this study and 77 previously collected L2 strains from the same population to reconstruct a maximum likelihood phylogenetic tree(*24, 25*). We showed that in this population there are three major clades: clade A consisting of the g2g-L2 isolates, and clade B and C (**Fig. 4A)**. We merged the WGS of these L2 clade isolates with 1,000 global L2 isolates that were previously published in 112 bioprojects (**Table S4**). We found the L2 strains from Peru formed subclades that were largely separated from other global L2 isolates (**Fig. S12**). This pattern of restriction suggests that *Mtb* L2 strains were introduced to Peru and then diversified locally.

**FIG 4.**
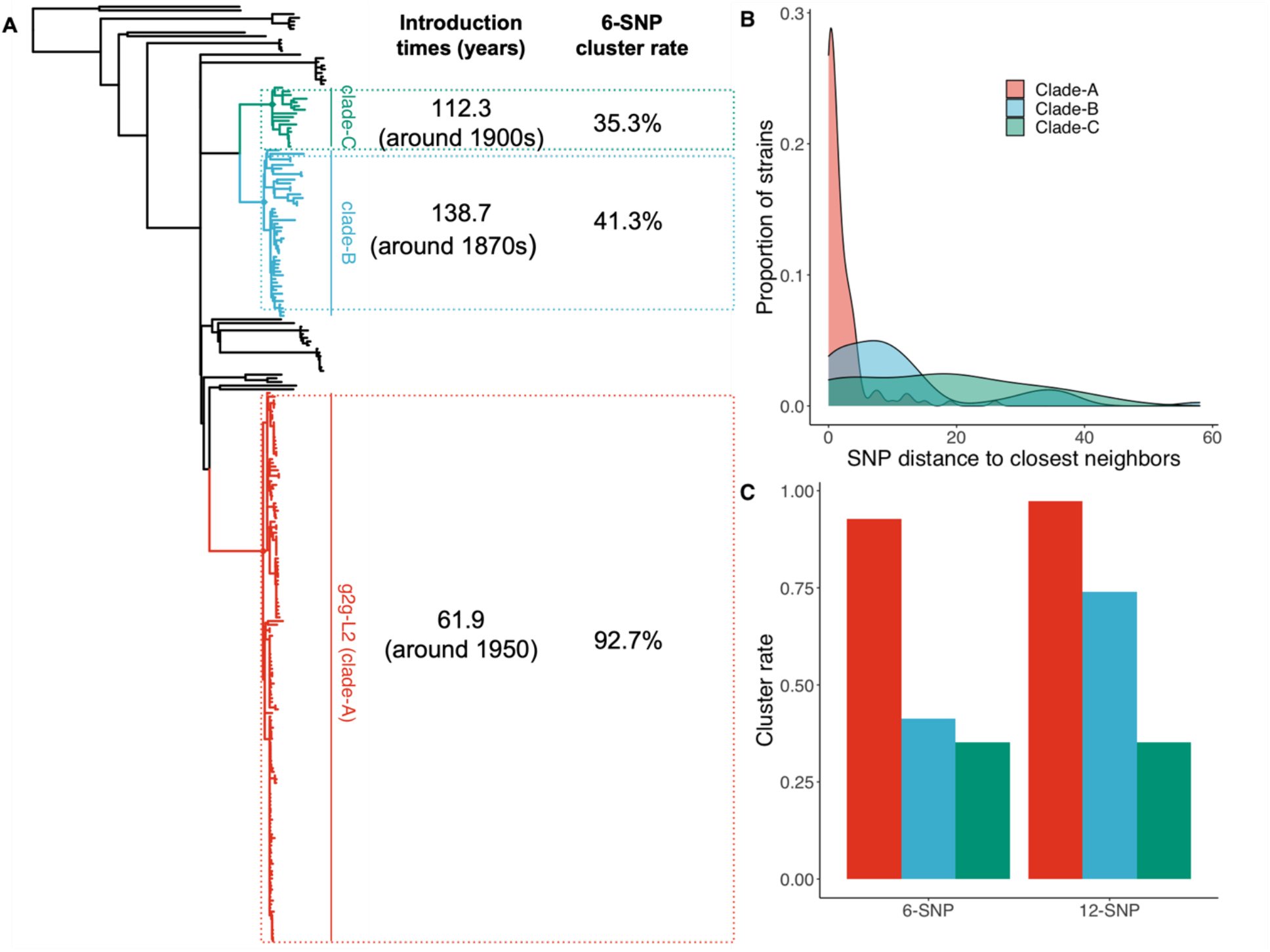
Genetic structure of the *Mtb* clade associated with the human *FLOT1* locus. (**A**) A phylogenetic tree of L2 constructed with 255 L2 Peruvian isolates. The g2g-L2 clade identified via the g2g analysis is highlighted in red (Clade-A), two other Peruvian subclades of L2 are highlighted in blue (Clade-B) and green (Clade-C) respectively. Estimated origin time (median value) for the ancestor strains and cluster rate when using 6-SNP distance of each marked L2 clade (Clade-A, B and C) are listed. (**B**) Histogram of pairwise minimum SNP distance to the closest neighbors within the marked clades. (**C**) Comparison of transmission cluster rate between the three marked clades when using 6-SNP (dark blue) and 12-SNP (light blue) distance as the threshold.

We next performed a Bayesian coalescent analysis to estimate the introduction times of these three L2 subclades(*26*). These results suggest that the g2g-L2 (clade A) was introduced to Peru more recently than the other two L2 subclades B and C (61.9 versus 138.7 and 112.3 years ago, **Fig. 4A**). We then compared the minimum SNP distance between these isolates and their closest neighbors in our WGS dataset to estimate an upper bound of the divergence time since the last transmission event for each TB case. The minimum SNP distances were significantly shorter for the g2g-L2 strains (median 1 SNPs) than isolates in clades B and C (median 8 and 18 SNPs for clade B and C respectively, P = 6.85 × 10^−13^ using the Kolmogorov-Smirnov test, **Fig. 4B**). To further compare the transmissibility to these clades, we assessed the frequency of L2 strains within our cohort. We expect that locally diversified strains which are introduced earlier would have a larger population size(*27, 28*). Instead, we found that the most recently introduced g2g-L2 strains have a larger than expected population size. Among 255 participants infected with L2 in our cohort, we observed 150 of 255 (58.8%) of them were infected by g2g-L2 strains (clade A), which was much higher than the number of infections caused by clade B or C (18.0% and 6.7%). These observations suggest that the g2g-L2 strains may have undergone a recent, rapid expansion.

Our findings raise the possibilities that g2g-L2 strains are more transmissible than other L2 clades or are more adept at causing disease in the Peruvian population starting in the 1950s after their introduction. To better understand the differences in the transmission dynamics among the three local clades, we explored the distribution of potential transmission clusters where two TB cases were considered to belong to the same transmission cluster if they were separated by fewer than 6 SNPs. We found among 150 participants infected with the g2g-L2 clade, 139 (92.7%) had a SNP distance smaller than 6-SNPs compared to closest neighbors, suggesting they are potentially linked by direct transmission. This cluster rate is much higher than that of other L2 strains collected from the same region (**Fig. S13**) (92.7% versus 40%, 42/105). We reached the same conclusion when using a less stringent 12-SNP cut-off (**Fig. 4C**). These data suggest that the g2g-L2 clade is associated with a higher level of recent transmission in the Peruvian population.

### Mtb g2g-L2 subclade displays differing redox metabolism from other L2 strains

To identify biological features that distinguish g2g-L2 strains, we phenotyped 23 g2g and 11 control L2 strains using a high throughput imaging-based phenotyping platform(*29*) (**Fig. 5A, Fig. S14**). We assessed seven functional phenotypes including cell morphology (length, width and area), DNA content with DAPI staining, the pattern and rate of peptidoglycan synthesis using NBD-amino-D alanine (NADA) incorporation, redox state using cyan fluorescent protein (CFP) autofluorescence, and intrabacterial lipid inclusions accumulation with Nile Red-staining. Among these phenotypes, we observed a significantly higher CFP signal in g2g compared to other L2 strains (Wilcoxon test, P =2.0 × 10^−4^, **Fig. 5B-C**), indicative of a shift in the redox state of F_420_ towards a more oxidized state. F_420_ is a redox active enzyme cofactor found in actinobacteria and methanogens that mediates low-reduction potential metabolic reactions(38). To determine if this functional difference in F_420_ results in biologically meaningful consequences, we assessed susceptibility to pretomanid, an antibiotic pro-drug that requires the reduced form of F_420_ to be activated(*30*). The g2g-L2 strains were more resistant to pretomanid, consistent with the measured shift towards oxidized F_420_ (Wilcox test, P = 0.032, **Fig. 5D**).

**FIG 5.**
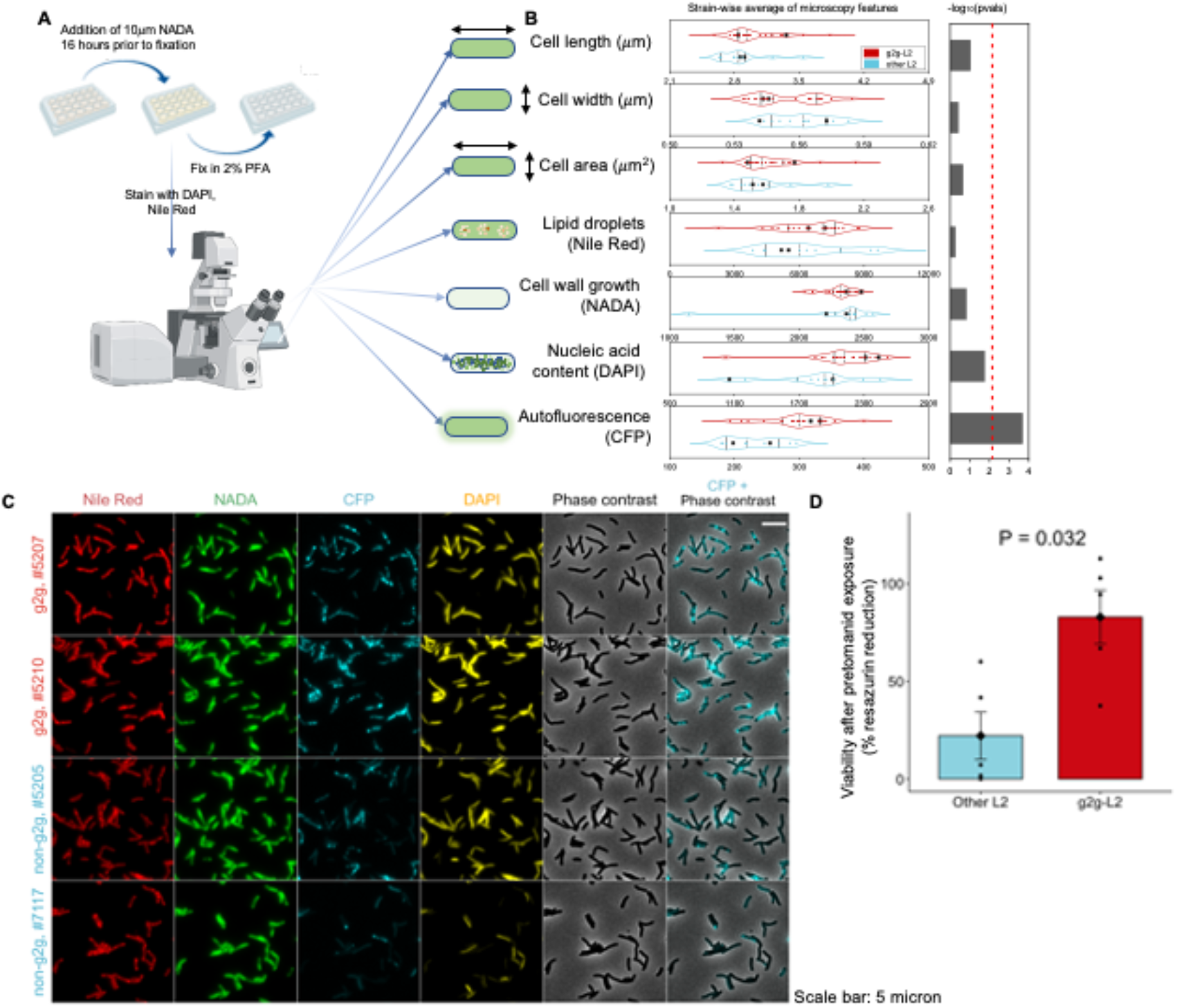
Functional characterization of g2g-L2 Mtb strains. (**A**) Schematic of the high throughput imaging-based phenotyping platform. 23 g2g-L2 and 11 other L2 *Mtb* strains were grown in 7H9 to a final OD600 of approximately 1.0. NADA (10uM) was added 16 hours prior to fixation with 2% paraformaldehyde. All samples were analyzed by microscopy to assess the population mean of cell length, cell width, cell area, DNA content using DAPI, intrabacterial lipid inclusions accumulation with Nile Red-staining, the pattern and rate of peptidoglycan synthesis using NADA incorporation, and redox state using CFP autofluorescence (oxidized form of F420 enzyme cofactor). (**B**) Violin plots for each phenotype described in (**A**), showing the distribution of each feature between the g2g and other L2 strains. Each point shows a representative sample for each individual strain; samples were assayed at minimum in duplicate (**Fig. S14**). The dotted lines inside each plot indicate the first and third quartiles; the dashed line indicates the median. Each point represents the average of two independent replicates per strain. A Wilcoxon test was performed to measure the difference between g2g and other L2 strains. The red dotted line indicates the Bonferroni corrected significance threshold after multiple testing (-log_10_(0.05/7)). (**C**) Representative images of Nile Red, NADA, CFP, and DAPI signals in two representative g2g-L2 strains and two non-g2g-L2 strains are shown (strains marked as enlarged black squares in (**B**)). (**D**) Based on a down-selection from the microscopy data, 5 g2g and 5 other L2 Mtb strains (two replicates per strain) were incubated with pretomanid (0.078uM) for 7 days. Resazurin was added to each well, and cultures were incubated an additional 24 hours. Variability after pretomanid exposure is measured by percent reduction in resazurin as calculated from drug-free controls. We used a Wilcoxon test for comparing the means of g2g and other L2 strains (P=0.032).

We examined putative *Mtb* genes that might be driving this functional phenotype. To this end, we identified nonsynonymous changes in 49 variants among 79 common *Mtb* variants defining g2g-L2 (**Table S2**). We found many of these occurred in genes that overlap with redox-related pathways (**Fig. S15**). In a gene set enrichment analysis, g2g-L2 was enriched for nonsynonymous mutations associated with flavin adenine dinucleotide (FAD) binding compared to all non-g2g-L2 nonsynonymous mutations (Fisher’s exact test, P = 0.008). FAD is a redox coenzyme of several important reactions in metabolism. Genes that overlap with the FAD binding pathway (GO:0050660) include *trxB2* (Thr2Asn), *fadE3* (Tyr59His), *fadE7* (Met364Ile), *fadE34* (Ala371Val), and *fadE35* (Asp417Asn). Three of these five mutations were predicted to attenuate protein function using SIFT(*31*) (SIFT score < 0.05, **Table S2**). Intriguingly we also observed a nonsynonymous mutation in *ctpC* (Asp633Asn) that has an attenuated function (SIFT score = 0.02). Previous study showed that cells lacking *ctpC* are both variable and sensitive to superoxide(*32*). Taken together, we found a cytological feature (oxidized form of F_420_) that can differentiate g2g-L2 strains from other co-circulating L2 strains, which can manifest as reduced susceptibility to the anti-TB drug pretomanid and concordant of mutations that accumulated by g2g-L2.

## Discussion

Previous work has shown that *Mtb* lineages are often restricted to particular geographical regions while other lineages are globally distributed, suggesting that *Mtb* can adapt to diverse natural environments and host populations(*12, 33*). In this work, we examined the hypothesis that genomic interactions between host and pathogen alleles can modify the risk of TB disease by performing a paired genome analysis using host and bacterial samples from a large longitudinal cohort of TB patients in Lima, Peru. We provided evidence (1) that a host regulatory variant in *FLOT1* gene is associated with a specific subset of *Mtb* strains; (2) that these strains form a subclade of the L2 lineage which is associated with specific human genetic variants; and (3) that this subclade, that we refer to as g2g-L2, has unique clinical and functional characteristics. Our unbiased g2g approach was essential to identify a human genetic association to a previously unrecognized L2 subclade.

Associations between host variants and specific pathogen genetic loci are not unique to TB, and have recently been found in many other infectious diseases such as human immunodeficiency virus, hepatitis C virus, Epstein-Barr virus and *Streptococcus pneumoniae* bacteria(*34*–*37*). Our study argues that inclusion of the genetic sequences of infectious agents can offer novel biological insights to future genetic studies of infectious diseases.

In the human host, we showed evidence of colocalization between the g2g signal and altered expression of the *FLOT1* gene in the lung and elsewhere. FLOT1 is a lipid-raft associated membrane protein that could alter the interaction with *Mtb* in multiple ways. FLOT1-dependent microdomains are present on the phagolysosome where they act as platforms for the assembly of NADPH oxidase complexes and vATPase(*18*). This function contributes to antifungal immunity, and *FLOT1* alleles are associated with invasive aspergillosis(*38*). *FLOT1* may play a similar role in *Mtb*-macrophage interactions. This protein also contributes to other signaling and cell migration pathways, raising the possibility that altered *FLOT1* expression could have additional effects on the host immune response to *Mtb*.

*FLOT1* interacts with g2g-L2, a unique *Mtb* L2 subclade that had undergone an expansion after its estimated introduction to Peru around the 1950s based on phylogenetic analysis. This may be due to its propensity to cause active TB. Consistent with this, we observed that the g2g-L2 subclade has a high transmission cluster rate among cases. Using unbiased phenotypic profiling *ex vivo* assays, we identified altered redox balance, and specifically altered F_420_ redox state, as a defining characteristic of g2g-L2 strains. Previous studies have shown that F_420_ protects *Mtb* to survive from oxidative and nitrosative stresses, and is also important for bacterial survival in the hypoxic environment characteristic of the human granuloma(*39*–*41*). Future studies will be required to determine whether the shift in redox balance is specific to F_420_ or whether it impacts other cofactor pools. Furthermore, we found g2g-L2 strains have acquired mutations in multiple genes that are critical for maintaining redox balance, including *trxB2* (Thr2Asn), which catalyzes thiol/disulfide exchange reactions to maintain cytoplasmic redox balance(*42*), and *ctpC* (Asp633Asn), which is required to metalate superoxide dismutase(*43*), and multiple FAD genes. This suggests that altered redox state is a primary phenotype for g2g-L2 strains. Importantly, we find that the shift towards oxidized F_420_ makes g2g-L2 strains intrinsically less susceptible to pretomanid, which requires reduced F_420_ to be effective. To our knowledge, this is the first evidence that host selection impacts the drug susceptibility of the bacterium, a concept that is likely to be an important paradigm for other new antimicrobials targeting bacterial energy metabolism.

Our work provides clear evidence of genetic interactions between variants in *Mtb* and human hosts that can determine the transmission dynamics and clinical outcome of tuberculosis. Further analysis is required to fully understand how the host *FLOT1* gene and g2g-L2 strain are mechanistically linked, and to establish their clinical relevance. Our results open the possibility that other *Mtb* strain specific host alleles are present, and may explain genetic differences driving TB susceptibility throughout the globe.

## Materials and Methods

### Ethics Statement

We recruited 1,632 subjects from a large catchment area of Lima, Peru that included 20 urban districts and ∼3.3 million residents to donate a blood sample for use in our study. We obtained written informed consent from all the participants. The study protocol was approved by the Institutional Review Board of Harvard School of Public Health and by the Research Ethics Committee of the National Institute of Health of Peru.

### Participant enrollment and follow-up

The study design and methods were previously described in detail(*24, 44*). Briefly, over the four-year period from 2009-2012, we identified patients ≥ 15 years of age who had received a diagnosis of pulmonary TB at any of 106 participating health centers. We confirmed the microbiological status of their disease with either a positive sputum smear or mycobacterial culture. We also recorded the index patients’ baseline smear status, HIV status, and drug-resistance profiles. Index cases with HIV infection or infected with multiple *Mtb* strains were excluded from the analyses.

### Host genotyping, quality control and imputation

DNA samples were genotyped using a customized genotyping array (LIMAArray) based on whole-exome sequencing data from 116 active TB cases to optimize the capture of genome-wide genetic variation in Peruvian individuals(*10*). We excluded samples that were missing more than 5% of the genotype data, had an excess of heterozygous genotypes, and/or duplicated with identity-by-state > 0.9. We also excluded variants with a call rate less than 95%, with duplicated position markers, minor allele frequency < 1%, and marked deviation from Hardy-Weinberg equilibrium (excluded if P < 10^−5^). After these quality control metrics, the genotype data were pre-phased using SHAPEIT2(*45*). We then used IMPUTE2(*46*) to impute genotypes at untyped genetic variants using the 1000 Genomes Project Phase 3(*20*) as the reference panel.

We imputed eight classical HLA genes HLA-*A, -B, -C*, -*DQA1, -DQB1, -DRB1, -DPA1*, and -*DPB1*, amino acids and intergenic variants using *HLA-TAPAS* using a multi-ancestry reference panel which contains data from 21,546 unrelated individuals(*47*).

### *Mycobacterium tuberculosis* variant calling and phylogeny construction

*Mtb* DNA was extracted from cryopreserved cultures of sputum samples. Each sample was thawed and subcultured on LJ and a big loop of colonies were lysed with lysozyme and proteinase K to obtain DNA using CTAB/Chloroform extraction and ethanol precipitation. Samples were sequenced on an Illumina HiSeq 2500 or 4000 sequencer yielding paired-end reads of length 125bp which were aligned to the reference assembly, H37Rv NC_000962.3 (GenBank accession CP003248). We called variants using Pilon (v1.22)(*48*). Genome coverage was assessed using Samtools (v1.10). We excluded isolates with evidence of mixed infection using the barcode method(*16*). We assigned a variant call as missing if the valid depth of coverage at a specific variant was less than 12 reads, if the mean read mapping quality at the site did not reach 10, or if none of the alternative alleles account for at least 85% of the valid coverage.

The phylogenetic tree was constructed based on the WGS *Mtb* alignment. Variants occurring in genes with repetitive elements such as transposases, proline-glutamate (PE) or proline-proline-glutamate (PPE)(*49*) were excluded to avoid any inaccuracies in the alignment. After applying these filters, 13,981 *Mtb* variants were used to conduct a genetic distance matrix. We built a phylogenetic tree using distance-based neighbor-joining coded in the *ape* package(*50*).

To characterize the Peruvian L2 *Mtb* strains in the context of global strains, we randomly selected 1,000 whole-genome sequenced L2 strains from published studies in 112 bioprojects to represent the major phylogenetic structures of L2. We used these global strains together with 255 L2 Peruvian strains to reconstruct the global L2 phylogenetic tree.

We estimated divergence times via BEAST v1.8.0(*26*), using an uncorrelated lognormal relaxed clock that allows for tree branches to evolve at different rates. The XML input file was modified to specify the number of invariant sites in the *Mtb* genomes. For the *Mtb* substitution rate, we used a normal distribution with a mean of 4.6 × 10^−8^ substitution per genome per site per year (3.0 × 10^−8^ to 6.2 × 10^−8^, 95% highest polar density interval), which was calibrated by ancient DNA samples(*51, 52*). An uncorrelated log-normal distribution was used for the substitution rate and a constant population size for the tree priors. We ran three chains of 5 × 10^−7^ generations and sampled every 10,000 generations to assure independent convergence of the chains; we discarded the first 10% as a burn-in. Convergence was assessed using Tracer (v1.7.0)(*53*), ensuring all relevant parameters reached an effective sample size > 100.

### Transmission cluster analysis

To define transmission clusters of the collected L2 *Mtb* isolates, we applied two SNP thresholds (6 and 12) to separate a patient isolate from that of at least one other patient in the cluster. The 6-SNP threshold was defined as the range of SNP distances between paired isolates of the same strain obtained at different times from relapsed TB patients(*54*). The 12-SNP threshold was a previously defined upper threshold of genomic relatedness noted within human hosts and between epidemiologically related human hosts(*55*).

### Genome-to-genome association analysis

To systematically test for interaction between human and *Mtb* genomic variants at the genome-wide level, we performed mixed logistic regression modeling using GEMMA(*56*) version 0.94.1 assuming an additive genetic model for each host and bacterial variant. For each bacterial variant *j* (*j* = 1,…, 1,267) and host variant *i* (*i* = 1,…, 676,110), we test

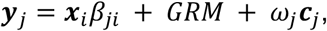

where *y*_*j*_ is an N-vector of binary labels indicating the absence or presence of a bacterial variant in the host samples i (n = 1,556); *x*_*i*_ is an N-vector of host genotypes; *β*_*ji*_ is the additive effect of the host variant i on the bacterial variant j; c_j_ is the j^th^ column of an N×C matrix of covariates (age and sex) including a column of 1s; and *ω*_*j*_ is a c-vector of the corresponding coefficients including the intercept. We used genetic relatedness matrix (GRM) as a random effect to correct for cryptic relatedness and population stratification between collected host samples. The GRM was obtained from an LD-pruned (r^2^ < 0.2) genotype with minor allele frequency >1%.

### Permutation Strategy

To determine an appropriate empirical genome-wide significance threshold for the g2g analysis accounting for population structure both in the *Mtb* and human genome, we conducted 200 simulations for each g2g analysis using a permutation procedure. To preserve the phenotype (defined by the *Mtb* variants) structure, for each interaction, we randomly permuted the presence/absence status for each of the 1,267 *Mtb* variants within the 1,556 individuals included in the study. In total, we ran 1,267 × 200 = 253,400 association tests to obtain an empirical genome-wide significance threshold.

We measured the distributions of the minimum p-values of the variants (P_min_) for each interaction of the permutation. We defined an empirical genome-wide significance threshold, - log_10_(P_sig_), as the 95th percentile (1-α) of -log_10_(P_min_).

### Colocalization analysis using eQTLs

We integrated our GWAS results with cis-eQTL data using a Bayesian method (coloc v3.2-1)(*57*). We evaluated whether the GWAS and eQTL associations best fit a model in which the associations are due to a single shared variant (summarized by the posterior probability) as opposed to different regulating variants. We used gene expression datasets from the eQTL catalog(*23*) release 4. We tested pairwise colocalization between 108 bulk RNA-sequencing expression datasets in 69 distinct cell and tissue types and GWAS from the most significant genome-to-genome pair (position 271640 as phenotype). We used GWAS and all variant-gene cis-eQTL associations tested in each tissue (including non-significant associations) of genes within a 800kb window around the top association host SNP (rs3130660). A posterior probability of colocalization >0.8 was considered to be a strong evidence for a causal gene.

### Bacterial strains and culture conditions

All 23 g2g and 11 non-g2g *Mtb* L2 strains were drug sensitive and HIV negative. They were grown shaking at 37°C. Cultures were grown in 7H9 media (Middlebrook 7H9 salts with 0.2% glycerol, 10% OADC [oleic acid, albumin, dextrose, catalase]). Cell density (OD600) was determined by a spectrophotometer.

### Microscopy imaging and analysis

*Mtb* cultures were grown to OD600 of approximately 1.0, then fixed with 2% paraformaldehyde (PFA) for 1 hour. The fluorescent D-amino acid NADA (3-[(7-Nitro-2,1,3-benzoxadiazol-4-yl)amino]-D alanine hydrochloride) was added at a final concentration of 1mM 16 hours prior to fixation. All samples were seeded onto molded 1.8% agarose in phosphate buffered saline (PBS) with DAPI (2.5μg/mL) and Nile red (0.1μg/mL). Samples were incubated at 37°C for one hour prior to imaging.

Samples were imaged with a Plan Apo 100× 1.45 NA objective using a Nikon Ti-E inverted, widefield microscope equipped with a Nikon Perfect Focus system with a Piezo Z drive motor, Andor Zyla sCMOS camera, and NIS Elements (v4.5). Semi-automated imaging was carried out using a customized Nikon JOBS script to locate imaging fields of interest, 24 images were taken for each strain. Cell segmentation and quantification was performed using our previously published pipeline, MOMIA(*29*).

### Minimum inhibitory concentration (MIC) determination

Pretomanid (PA-824) MIC value for each strain was determined following the microplate-based Alamar Blue assay method as previously described(*58*). 96-well plates containing two-fold serial dilutions of the antibiotic Pretomanid (PA-824) in 7H9 media were prepared, creating a concentration gradient from 40 to 0.039 µM. A drug-free control well was included. All *Mtb* strains were grown to mid-log phase and diluted to OD600 = 0.003 into each well of the prepared plates. Plates were incubated at 37°C without shaking in sealed plates. After 7 days, a freshly prepared mixture of 0.15mg/ml resazurin (Sigma Aldrich) diluted in PBS and 20%

Tween-80 was added to each well. Plates were incubated for 24 hours, after which absorbance was read at 570 nm, with a reference wavelength of 600 nm. Percent reduction of resazurin was normalized for each strain by subtracting the OD570 of the minimum value wells (40µM drug condition) and scaling by the reduction measurement of the no-drug control. Data are representative of two independent replicates per strain.

### Pathway analysis of interacting Mtb genes

We used Variant Effect Predictor(*59*) (v.105) to first annotate the *Mtb* genome. Among 79 highly correlated (Pearson r^2^>0.8) common *Mtb* variants that are associated with the human genome, 49 were annotated as nonsynonymous (missense) variants. We defined *Mtb* genes that have one of these 49 missense variants as g2g-L2 genes. We used the DAVID(*60*) Functional Annotation Clustering function to group g2g-L2 genes into a similar functional network. We used PANTHER(*61*) to test for functional overlaps with known pathways. We calculated a Fisher’s exact p-value by constructing a 2×2 contingency table with g2g-L2 genes (n = 48) and all other *Mtb* genes with a common missense mutation in the g2g analysis (n = 898) overlapping the flavin adenine dinucleotide binding pathway included in the gene ontology category (GO:0050660).

## Data Availability

All data for generating the figures presented in the manuscript are available at https://github.com/immunogenomics/TB-g2g.
Human genotyping data are available through dbGAP, under accession number phs002025.v1.p1.

## Author contributions

S.R. and M.B.M. conceptualized and designed the study. Y.L. and S.R. designed the statistical and computational strategy and analyzed the data. C.H. performed phylogenetic analysis of the *Mtb* genome. Q.L. and S.M.F. designed and performed phylogenomic analysis of all L2 strains. Q.L. N.H., J.Z. and S.M.F. designed and performed *ex vivo* microscopy assay of L2 strains. X.L. annotated the *Mtb* genome. T.A. performed functional annotation of the host associated alleles. S.A. performed ancestry inference of the host genome. K.I. performed colocalization analysis of the blueprint data. D.B.M. provided feedback on data interpretation. R.C., L.L and M.B.M. collected and curated the clinical and genomic data. Y.L. and S.R. wrote the initial manuscript. All authors contributed to the writing and editing of the final manuscript.

## Data and materials availability

All data for generating the figures presented in the manuscript are available at https://github.com/immunogenomics/TB-g2g.

Human genotyping data are available through dbGAP, under accession number phs002025.v1.p1.

## References

1. C. Tian, B. S. Hromatka, A. K. Kiefer, N. Eriksson, S. M. Noble, J. Y. Tung, D. A. Hinds, Genome-wide association and HLA region fine-mapping studies identify susceptibility loci for multiple common infections. Nat. Commun. 8, 599 (2017).

2. S. Sakaue, M. Kanai, Y. Tanigawa, J. Karjalainen, M. Kurki, S. Koshiba, A. Narita, T. Konuma, K. Yamamoto, M. Akiyama, K. Ishigaki, A. Suzuki, K. Suzuki, W. Obara, K. Yamaji, K. Takahashi, S. Asai, Y. Takahashi, T. Suzuki, N. Shinozaki, H. Yamaguchi, S. Minami, S. Murayama, K. Yoshimori, S. Nagayama, D. Obata, M. Higashiyama, A. Masumoto, Y. Koretsune, FinnGen, K. Ito, C. Terao, T. Yamauchi, I. Komuro, T. Kadowaki, G. Tamiya, M. Yamamoto, Y. Nakamura, M. Kubo, Y. Murakami, K. Yamamoto, Y. Kamatani, A. Palotie, M. A. Rivas, M. J. Daly, K. Matsuda, Y. Okada, A cross-population atlas of genetic associations for 220 human phenotypes. Nat. Genet. 53, 1415–1424 (2021).

3. R. M. G. J. Houben, P. J. Dodd, The Global Burden of Latent Tuberculosis Infection: A Reestimation Using Mathematical Modelling. PLoS Med. 13, e1002152 (2016).

4. W. H. Organization, Others, Global tuberculosis report 2018. 2018. Geneva: World Health Organization (2019).

5. T. Thye, F. O. Vannberg, S. H. Wong, E. Owusu-Dabo, I. Osei, J. Gyapong, G. Sirugo, F. Sisay-Joof, A. Enimil, M. A. Chinbuah, S. Floyd, D. K. Warndorff, L. Sichali, S. Malema, A. C. Crampin, B. Ngwira, Y. Y. Teo, K. Small, K. Rockett, D. Kwiatkowski, P. E. Fine, P. C. Hill, M. Newport, C. Lienhardt, R. A. Adegbola, T. Corrah, A. Ziegler, African TB Genetics Consortium, Wellcome Trust Case Control Consortium, A. P. Morris, C. G. Meyer, R. D. Horstmann, A. V. S. Hill, Genome-wide association analyses identifies a susceptibility locus for tuberculosis on chromosome 18q11.2. Nat. Genet. 42, 739–741 (2010).

6. S. Mahasirimongkol, H. Yanai, T. Mushiroda, W. Promphittayarat, S. Wattanapokayakit, J. Phromjai, R. Yuliwulandari, N. Wichukchinda, A. Yowang, N. Yamada, P. Kantipong, A. Takahashi, M. Kubo, P. Sawanpanyalert, N. Kamatani, Y. Nakamura, K. Tokunaga, Genome-wide association studies of tuberculosis in Asians identify distinct at-risk locus for young tuberculosis. J. Hum. Genet. 57, 363–367 (2012).

7. T. Thye, E. Owusu-Dabo, F. O. Vannberg, R. van Crevel, J. Curtis, E. Sahiratmadja, Y. Balabanova, C. Ehmen, B. Muntau, G. Ruge, J. Sievertsen, J. Gyapong, V. Nikolayevskyy, P. C. Hill, G. Sirugo, F. Drobniewski, E. van de Vosse, M. Newport, B. Alisjahbana, S. Nejentsev, T. H. M. Ottenhoff, A. V. S. Hill, R. D. Horstmann, C. G. Meyer, Common variants at 11p13 are associated with susceptibility to tuberculosis. Nat. Genet. 44, 257–259 (2012).

8. E. R. Chimusa, N. Zaitlen, M. Daya, M. Möller, P. D. van Helden, N. J. Mulder, A. L. Price, E. G. Hoal, Genome-wide association study of ancestry-specific TB risk in the South African Coloured population. Hum. Mol. Genet. 23, 796–809 (2014).

9. J. Curtis, Y. Luo, H. L. Zenner, D. Cuchet-Lourenço, C. Wu, K. Lo, M. Maes, A. Alisaac, E. Stebbings, J. Z. Liu, L. Kopanitsa, O. Ignatyeva, Y. Balabanova, V. Nikolayevskyy, I. Baessmann, T. Thye, C. G. Meyer, P. Nürnberg, R. D. Horstmann, F. Drobniewski, V. Plagnol, J. C. Barrett, S. Nejentsev, Susceptibility to tuberculosis is associated with variants in the ASAP1 gene encoding a regulator of dendritic cell migration. Nat. Genet. 47, 523–527 (2015).

10. Y. Luo, S. Suliman, S. Asgari, T. Amariuta, Y. Baglaenko, M. MartÍnez-Bonet, K. Ishigaki, M. Gutierrez-Arcelus, R. Calderon, L. Lecca, S. R. León, J. Jimenez, R. Yataco, C. Contreras, J. T. Galea, M. Becerra, S. Nejentsev, P. A. Nigrovic, D. B. Moody, M. B. Murray, S. Raychaudhuri, Early progression to active tuberculosis is a highly heritable trait driven by 3q23 in Peruvians. Nat. Commun. 10, 3765 (2019).

11. S. Gagneux, Host–pathogen coevolution in human tuberculosis. Philos. Trans. R. Soc. Lond. B Biol. Sci. 367, 850–859 (2012).

12. I. Comas, M. Coscolla, T. Luo, S. Borrell, K. E. Holt, M. Kato-Maeda, J. Parkhill, B. Malla, Berg, G. Thwaites, D. Yeboah-Manu, G. Bothamley, J. Mei, L. Wei, S. Bentley, S. R. Harris, S. Niemann, R. Diel, A. Aseffa, Q. Gao, D. Young, S. Gagneux, Out-of-Africa migration and Neolithic coexpansion of Mycobacterium tuberculosis with modern humans. Nat. Genet. 45, 1176–1182 (2013).

13. M. L. McHenry, J. Bartlett, R. P. Igo Jr, E. M. Wampande, P. Benchek, H. Mayanja-Kizza, K. Fluegge, N. B. Hall, S. Gagneux, S. A. Tishkoff, C. Wejse, G. Sirugo, W. H. Boom, M. Joloba, S. M. Williams, C. M. Stein, Interaction between host genes and Mycobacterium tuberculosis lineage can affect tuberculosis severity: Evidence for coevolution? PLoS Genet. 16, e1008728 (2020).

14. Y. Omae, L. Toyo-Oka, H. Yanai, S. Nedsuwan, S. Wattanapokayakit, N. Satproedprai, N. Smittipat, P. Palittapongarnpim, P. Sawanpanyalert, W. Inunchot, E. Pasomsub, N. Wichukchinda, T. Mushiroda, M. Kubo, K. Tokunaga, S. Mahasirimongkol, Pathogen lineage-based genome-wide association study identified CD53 as susceptible locus in tuberculosis. J. Hum. Genet. 62, 1015–1022 (2017).

15. World Health Organization, Global Tuberculosis Report 2019 (World Health Organization, 2019; https://play.google.com/store/books/details?id=s0vszAEACAAJ).

16. F. Coll, R. McNerney, J. A. Guerra-Assunção, J. R. Glynn, J. Perdigão, M. Viveiros, I. Portugal, A. Pain, N. Martin, T. G. Clark, A robust SNP barcode for typing Mycobacterium tuberculosis complex strains. Nat. Commun. 5, 4812 (2014).

17. GeneCards Human Gene Database, FLOT1 Gene - GeneCards, (available at https://www.genecards.org/cgi-bin/carddisp.pl?gene=FLOT1).

18. J. F. Dermine, S. Duclos, J. Garin, F. St-Louis, S. Rea, R. G. Parton, M. Desjardins, Flotillin-1-enriched lipid raft domains accumulate on maturing phagosomes. J. Biol. Chem. 276, 18507–18512 (2001).

19. The GTEx Consortium, The GTEx Consortium atlas of genetic regulatory effects across human tissues. Science. 369, 1318–1330 (2020).

20. 1000 Genomes Project Consortium, A. Auton, L. D. Brooks, R. M. Durbin, E. P. Garrison, H. M. Kang, J. O. Korbel, J. L. Marchini, S. McCarthy, G. A. McVean, G. R. Abecasis, A global reference for human genetic variation. Nature. 526, 68–74 (2015).

21. C. Wallace, Statistical testing of shared genetic control for potentially related traits. Genet. Epidemiol. 37, 802–813 (2013).

22. N. R. Gay, M. Gloudemans, M. L. Antonio, N. S. Abell, B. Balliu, Y. Park, A. R. Martin, S. Musharoff, A. S. Rao, F. Aguet, A. N. Barbeira, R. Bonazzola, F. Hormozdiari, GTEx Consortium, K. G. Ardlie, C. D. Brown, H. K. Im, T. Lappalainen, X. Wen, S. B. Montgomery, Impact of admixture and ancestry on eQTL analysis and GWAS colocalization in GTEx. Genome Biol. 21, 233 (2020).

23. N. Kerimov, J. D. Hayhurst, K. Peikova, J. R. Manning, P. Walter, L. Kolberg, M. SamoviČa, M. P. Sakthivel, I. Kuzmin, S. J. Trevanion, T. Burdett, S. Jupp, H. Parkinson, I. Papatheodorou, A. D. Yates, D. R. Zerbino, K. Alasoo, A compendium of uniformly processed human gene expression and splicing quantitative trait loci. Nat. Genet. 53, 1290–1299 (2021).

24. M. C. Becerra, C.-C. Huang, L. Lecca, J. Bayona, C. Contreras, R. Calderon, R. Yataco, J. Galea, Z. Zhang, S. Atwood, T. Cohen, C. D. Mitnick, P. Farmer, M. Murray, Transmissibility and potential for disease progression of drug resistant Mycobacterium tuberculosis: prospective cohort study. BMJ. 367, l5894 (2019).

25. M. R. Farhat, L. Freschi, R. Calderon, T. Ioerger, M. Snyder, C. J. Meehan, B. de Jong, L. Rigouts, A. Sloutsky, D. Kaur, S. Sunyaev, D. van Soolingen, J. Shendure, J. Sacchettini, M. Murray, GWAS for quantitative resistance phenotypes in Mycobacterium tuberculosis reveals resistance genes and regulatory regions. Nat. Commun. 10, 2128 (2019).

26. A. J. Drummond, M. A. Suchard, D. Xie, A. Rambaut, Bayesian phylogenetics with BEAUti and the BEAST 1.7. Mol. Biol. Evol. 29, 1969–1973 (2012).

27. Q. Liu, A. Ma, L. Wei, Y. Pang, B. Wu, T. Luo, Y. Zhou, H.-X. Zheng, Q. Jiang, M. Gan, T. Zuo, M. Liu, C. Yang, L. Jin, I. Comas, S. Gagneux, Y. Zhao, C. S. Pepperell, Q. Gao, China’s tuberculosis epidemic stems from historical expansion of four strains of Mycobacterium tuberculosis. Nat Ecol Evol. 2, 1982–1992 (2018).

28. Q. Liu, H. Liu, L. Shi, M. Gan, X. Zhao, L.-D. Lyu, H. E. Takiff, K. Wan, Q. Gao, Local adaptation of Mycobacterium tuberculosis on the Tibetan Plateau. Proc. Natl. Acad. Sci. U. S. A. 118 (2021), doi:10.1073/pnas.2017831118.

29. J. Zhu, I. D. Wolf, C. L. Dulberger, H. I. Won, J. C. Kester, J. A. Judd, S. E. Wirth, R. R. Clark, Y. Li, Y. Luo, T. A. Gray, J. T. Wade, K. M. Derbyshire, S. M. Fortune, E. J. Rubin, Spatiotemporal localization of proteins in mycobacteria. Cell Rep. 37, 110154 (2021).

30. M. R. Hasan, M. Rahman, S. Jaques, E. Purwantini, L. Daniels, Glucose 6-phosphate accumulation in mycobacteria: implications for a novel F420-dependent anti-oxidant defense system. J. Biol. Chem. 285, 19135–19144 (2010).

31. R. Vaser, S. Adusumalli, S. N. Leng, M. Sikic, P. C. Ng, SIFT missense predictions for genomes. Nat. Protoc. 11, 1–9 (2016).

32. T. Padilla-Benavides, J. E. Long, D. Raimunda, C. M. Sassetti, J. M. Argüello, A Novel P1B-type Mn2 -transporting ATPase Is Required for Secreted Protein Metallation in Mycobacteria. Journal of Biological Chemistry. 288 (2013), pp. 11334–11347.

33. D. Stucki, D. Brites, L. Jeljeli, M. Coscolla, Q. Liu, A. Trauner, L. Fenner, L. Rutaihwa, S. Borrell, T. Luo, Q. Gao, M. Kato-Maeda, M. Ballif, M. Egger, R. Macedo, H. Mardassi, M. Moreno, G. T. Vilanova, J. Fyfe, M. Globan, J. Thomas, F. Jamieson, J. L. Guthrie, A. Asante-Poku, D. Yeboah-Manu, E. Wampande, W. Ssengooba, M. Joloba, W. Henry Boom, I. Basu, J. Bower, M. Saraiva, S. E. G. Vasconcellos, P. Suffys, A. Koch, R. Wilkinson, L. Gail-Bekker, B. Malla, S. D. Ley, H.-P. Beck, B. C. de Jong, K. Toit, E. Sanchez-Padilla, M. Bonnet, A. Gil-Brusola, M. Frank, V. N. Penlap Beng, K. Eisenach, I. Alani, P. W. Ndung’u, G. Revathi, F. Gehre, S. Akter, F. Ntoumi, L. Stewart-Isherwood, N. E. Ntinginya, A. Rachow, M. Hoelscher, D. M. Cirillo, G. Skenders, S. Hoffner, D. Bakonyte, P. Stakenas, R. Diel, V. Crudu, O. Moldovan, S. Al-Hajoj, L. Otero, F. Barletta, E. Jane Carter, L. Diero, P. Supply, I. Comas, S. Niemann, S. Gagneux, Mycobacterium tuberculosis lineage 4 comprises globally distributed and geographically restricted sublineages. Nature Genetics. 48 (2016), pp. 1535–1543.

34. I. Bartha, J. M. Carlson, C. J. Brumme, P. J. McLaren, Z. L. Brumme, M. John, D. W. Haas, J. Martinez-Picado, J. Dalmau, C. López-GalÍndez, C. Casado, A. Rauch, H. F. Günthard, E. Bernasconi, P. Vernazza, T. Klimkait, S. Yerly, S. J. O’Brien, J. Listgarten, N. Pfeifer, C. Lippert, N. Fusi, Z. Kutalik, T. M. Allen, V. Müller, P. R. Harrigan, D. Heckerman, A. Telenti, J. Fellay, A genome-to-genome analysis of associations between human genetic variation, HIV-1 sequence diversity, and viral control. Elife. 2, e01123 (2013).

35. M. A. Ansari, V. Pedergnana, C. L C Ip, A. Magri, A. Von Delft, D. Bonsall, N. Chaturvedi, I. Bartha, D. Smith, G. Nicholson, G. McVean, A. Trebes, P. Piazza, J. Fellay, G. Cooke, G. R. Foster, STOP-HCV Consortium, E. Hudson, J. McLauchlan, P. Simmonds, R. Bowden, P. Klenerman, E. Barnes, C. C. A. Spencer, Genome-to-genome analysis highlights the effect of the human innate and adaptive immune systems on the hepatitis C virus. Nat. Genet. 49, 666–673 (2017).

36. J. A. Lees, B. Ferwerda, P. H. C. Kremer, N. E. Wheeler, M. V. Serón, N. J. Croucher, R. A. Gladstone, H. J. Bootsma, N. Y. Rots, A. J. Wijmega-Monsuur, E. A. M. Sanders, K. Trzciński, A. L. Wyllie, A. H. Zwinderman, L. H. van den Berg, W. van Rheenen, J. H. Veldink, Z. B. Harboe, L. F. Lundbo, L. C. P. G. M. de Groot, N. M. van Schoor, N. van der Velde, L. H. Ängquist, T. I. A. Sørensen, E. A. Nohr, A. J. Mentzer, T. C. Mills, J. C. Knight, M. du Plessis, S. Nzenze, J. N. Weiser, J. Parkhill, S. Madhi, T. Benfield, A. von Gottberg, A. van der Ende, M. C. Brouwer, J. C. Barrett, S. D. Bentley, D. van de Beek, Joint sequencing of human and pathogen genomes reveals the genetics of pneumococcal meningitis. Nat. Commun. 10, 2176 (2019).

37. S. Rüeger, C. Hammer, A. Loetscher, P. J. McLaren, D. Lawless, O. Naret, D. P. Depledge, Morfopoulou, J. Breuer, E. Zdobnov, J. Fellay, Swiss HIV Cohort Study, The influence of human genetic variation on Epstein-Barr virus sequence diversity. Sci. Rep. 11, 4586 (2021).

38. F. Schmidt, A. Thywißen, M. Goldmann, C. Cunha, Z. Cseresnyés, H. Schmidt, M. Rafiq, S. Galiani, M. H. Gräler, G. Chamilos, J. F. Lacerda, A. Campos Jr, C. Eggeling, M. T. Figge, Heinekamp, S. G. Filler, A. Carvalho, A. A. Brakhage, Flotillin-Dependent Membrane Microdomains Are Required for Functional Phagolysosomes against Fungal Infections. Cell Rep. 32, 108017 (2020).

39. E. Purwantini, B. Mukhopadhyay, Conversion of NO2 to NO by reduced coenzyme F420 protects mycobacteria from nitrosative damage. Proc. Natl. Acad. Sci. U. S. A. 106, 6333–6338 (2009).

40. M. Gurumurthy, M. Rao, T. Mukherjee, S. P. S. Rao, H. I. Boshoff, T. Dick, C. E. Barry 3rd, H. Manjunatha, A novel F(420) -dependent anti-oxidant mechanism protects Mycobacterium tuberculosis against oxidative stress and bactericidal agents. Mol. Microbiol. 87, 744–755 (2013).

41. T. Jirapanjawat, B. Ney, M. C. Taylor, A. C. Warden, S. Afroze, R. J. Russell, B. M. Lee, C. J. Jackson, J. G. Oakeshott, G. Pandey, C. Greening, The redox cofactor F 420 protects mycobacteria from diverse antimicrobial compounds and mediates a reductive detoxification system. Appl. Environ. Microbiol. 82, 6810–6818 (2016).

42. M. Akif, G. Khare, A. K. Tyagi, S. C. Mande, A. A. Sardesai, Functional studies of multiple thioredoxins from Mycobacterium tuberculosis. J. Bacteriol. 190, 7087–7095 (2008).

43. S. Nambi, J. E. Long, B. B. Mishra, R. Baker, K. C. Murphy, A. J. Olive, H. P. Nguyen, S. A. Shaffer, C. M. Sassetti, The Oxidative Stress Network of Mycobacterium tuberculosis Reveals Coordination between Radical Detoxification Systems. Cell Host Microbe. 17, 829–837 (2015).

44. C.-C. Huang, A. L. Chu, M. C. Becerra, J. T. Galea, R. Calderón, C. Contreras, R. Yataco, Z. Zhang, L. Lecca, M. B. Murray, Mycobacterium tuberculosis Beijing Lineage and Risk for Tuberculosis in Child Household Contacts, Peru. Emerg. Infect. Dis. 26, 568–578 (2020).

45. J. O’Connell, D. Gurdasani, O. Delaneau, N. Pirastu, S. Ulivi, M. Cocca, M. Traglia, J. Huang, J. E. Huffman, I. Rudan, R. McQuillan, R. M. Fraser, H. Campbell, O. Polasek, G. Asiki, K. Ekoru, C. Hayward, A. F. Wright, V. Vitart, P. Navarro, J.-F. Zagury, J. F. Wilson, D. Toniolo, P. Gasparini, N. Soranzo, M. S. Sandhu, J. Marchini, A general approach for haplotype phasing across the full spectrum of relatedness. PLoS Genet. 10, e1004234 (2014).

46. B. Howie, C. Fuchsberger, M. Stephens, J. Marchini, G. R. Abecasis, Fast and accurate genotype imputation in genome-wide association studies through pre-phasing. Nat. Genet. 44, 955–959 (2012).

47. Luo, Yang and Kanai, Masahiro and Choi, Wanson and Li, Xinyi and Yamamoto, Kenichi and Ogawa, Kotaro and Gutierrez-Arcelus, Maria and Gregersen, Peter K and Stuart, Philip E and Elder, James T and Fellay, Jacques and Carrington, Mary and Haas, David W and Guo, Xiuqing and Palmer, Nicholette D and Chen, Yii-Der Ida and Rotter, Jerome I and Taylor, Kent D and Rich, Steve and Correa, Adolfo and Wilson, James G and Kathiresan, Sekar and Cho, Michael and Metspalu, Andres and Esko, Tonu and Okada, Yukinori and Han, Buhm and, and Mclaren, Paul J and Raychaudhuri, Soumya, A high-resolution HLA reference panel capturing global population diversity enables multi-ethnic fine-mapping in HIV host response. medRxiv (2020) (available at https://www.medrxiv.org/content/10.1101/2020.07.16.20155606v1).

48. B. J. Walker, T. Abeel, T. Shea, M. Priest, A. Abouelliel, S. Sakthikumar, C. A. Cuomo, Q. Zeng, J. Wortman, S. K. Young, A. M. Earl, Pilon: an integrated tool for comprehensive microbial variant detection and genome assembly improvement. PLoS One. 9, e112963 (2014).

49. J. E. Phelan, F. Coll, I. Bergval, R. M. Anthony, R. Warren, S. L. Sampson, N. C. G. van Pittius, J. R. Glynn, A. C. Crampin, A. Alves, T. B. Bessa, S. Campino, K. Dheda, L. Grandjean, R. Hasan, Z. Hasan, A. Miranda, D. Moore, S. Panaiotov, J. Perdigao, I. Portugal, P. Sheen, E. de Oliveira Sousa, E. M. Streicher, P. D. van Helden, M. Viveiros, M. L. Hibberd, A. Pain, R. McNerney, T. G. Clark, Recombination in pe/ppe genes contributes to genetic variation in Mycobacterium tuberculosis lineages. BMC Genomics. 17 (2016),, doi:10.1186/s12864-016-2467-y.

50. E. Paradis, K. Schliep, ape 5.0: an environment for modern phylogenetics and evolutionary analyses in R. Bioinformatics. 35, 526–528 (2019).

51. K. I. Bos, K. M. Harkins, A. Herbig, M. Coscolla, N. Weber, I. Comas, S. A. Forrest, J. M. Bryant, S. R. Harris, V. J. Schuenemann, T. J. Campbell, K. Majander, A. K. Wilbur, R. A. Guichon, D. L. Wolfe Steadman, D. C. Cook, S. Niemann, M. A. Behr, M. Zumarraga, R. Bastida, D. Huson, K. Nieselt, D. Young, J. Parkhill, J. E. Buikstra, S. Gagneux, A. C. Stone, J. Krause, Pre-Columbian mycobacterial genomes reveal seals as a source of New World human tuberculosis. Nature. 514, 494–497 (2014).

52. S. Sabin, A. Herbig, Å. J. Vågene, T. Ahlström, G. Bozovic, C. Arcini, D. Kühnert, K. I. Bos, A seventeenth-century Mycobacterium tuberculosis genome supports a Neolithic emergence of the Mycobacterium tuberculosis complex. Genome Biology. 21 (2020),, doi:10.1186/s13059-020-02112-1.

53. A. Rambaut, A. J. Drummond, D. Xie, G. Baele, M. A. Suchard, Posterior Summarization in Bayesian Phylogenetics Using Tracer 1.7. Systematic Biology. 67 (2018), pp. 901–904.

54. J. M. Bryant, S. R. Harris, J. Parkhill, R. Dawson, A. H. Diacon, P. van Helden, A. Pym, A. A. Mahayiddin, C. Chuchottaworn, I. M. Sanne, C. Louw, M. J. Boeree, M. Hoelscher, T. D. McHugh, A. L. C. Bateson, R. D. Hunt, S. Mwaigwisya, L. Wright, S. H. Gillespie, S. D. Bentley, Whole-genome sequencing to establish relapse or re-infection with Mycobacterium tuberculosis: a retrospective observational study. Lancet Respir Med. 1, 786–792 (2013).

55. T. M. Walker, C. L. C. Ip, R. H. Harrell, J. T. Evans, G. Kapatai, M. J. Dedicoat, D. W. Eyre, D. J. Wilson, P. M. Hawkey, D. W. Crook, J. Parkhill, D. Harris, A. S. Walker, R. Bowden, P. Monk, E. G. Smith, T. E. A. Peto, Whole-genome sequencing to delineate Mycobacterium tuberculosis outbreaks: a retrospective observational study. Lancet Infect. Dis. 13, 137–146 (2013).

56. H. Chen, C. Wang, M. P. Conomos, A. M. Stilp, Z. Li, T. Sofer, A. A. Szpiro, W. Chen, J. M. Brehm, J. C. Celedón, S. Redline, G. J. Papanicolaou, T. A. Thornton, C. C. Laurie, K. Rice, X. Lin, Control for Population Structure and Relatedness for Binary Traits in Genetic Association Studies via Logistic Mixed Models. Am. J. Hum. Genet. 98, 653–666 (2016).

57. C. Giambartolomei, D. Vukcevic, E. E. Schadt, L. Franke, A. D. Hingorani, C. Wallace, V. Plagnol, Bayesian test for colocalisation between pairs of genetic association studies using summary statistics. PLoS Genet. 10, e1004383 (2014).

58. S. G. Franzblau, R. S. Witzig, J. C. McLaughlin, P. Torres, G. Madico, A. Hernandez, M. T. Degnan, M. B. Cook, V. K. Quenzer, R. M. Ferguson, R. H. Gilman, Rapid, low-technology MIC determination with clinical Mycobacterium tuberculosis isolates by using the microplate Alamar Blue assay. J. Clin. Microbiol. 36, 362–366 (1998).

59. W. McLaren, L. Gil, S. E. Hunt, H. S. Riat, G. R. S. Ritchie, A. Thormann, P. Flicek, F. Cunningham, The Ensembl Variant Effect Predictor. Genome Biol. 17, 122 (2016).

60. X. Jiao, B. T. Sherman, D. W. Huang, R. Stephens, M. W. Baseler, H. C. Lane, R. A. Lempicki, DAVID-WS: a stateful web service to facilitate gene/protein list analysis. Bioinformatics. 28, 1805–1806 (2012).

61. H. Mi, D. Ebert, A. Muruganujan, C. Mills, L.-P. Albou, T. Mushayamaha, P. D. Thomas, PANTHER version 16: a revised family classification, tree-based classification tool, enhancer regions and extensive API. Nucleic Acids Res. 49, D394–D403 (2021).

