## Supplementary_Figures for "A *FLOT1* host regulatory allele is associated with a recently expanded *Mtb* clade in patients with tuberculosis"

**Figure S1. Histogram of quantile-quantile plots for test of association between each pair of human host and *Mtb* variants.** A histogram summarizing all genomic inflation factors over 1,267 genome-wide association studies. The distribution ranges between 0.97 and 1.07 with median 1.01 (red vertical line).

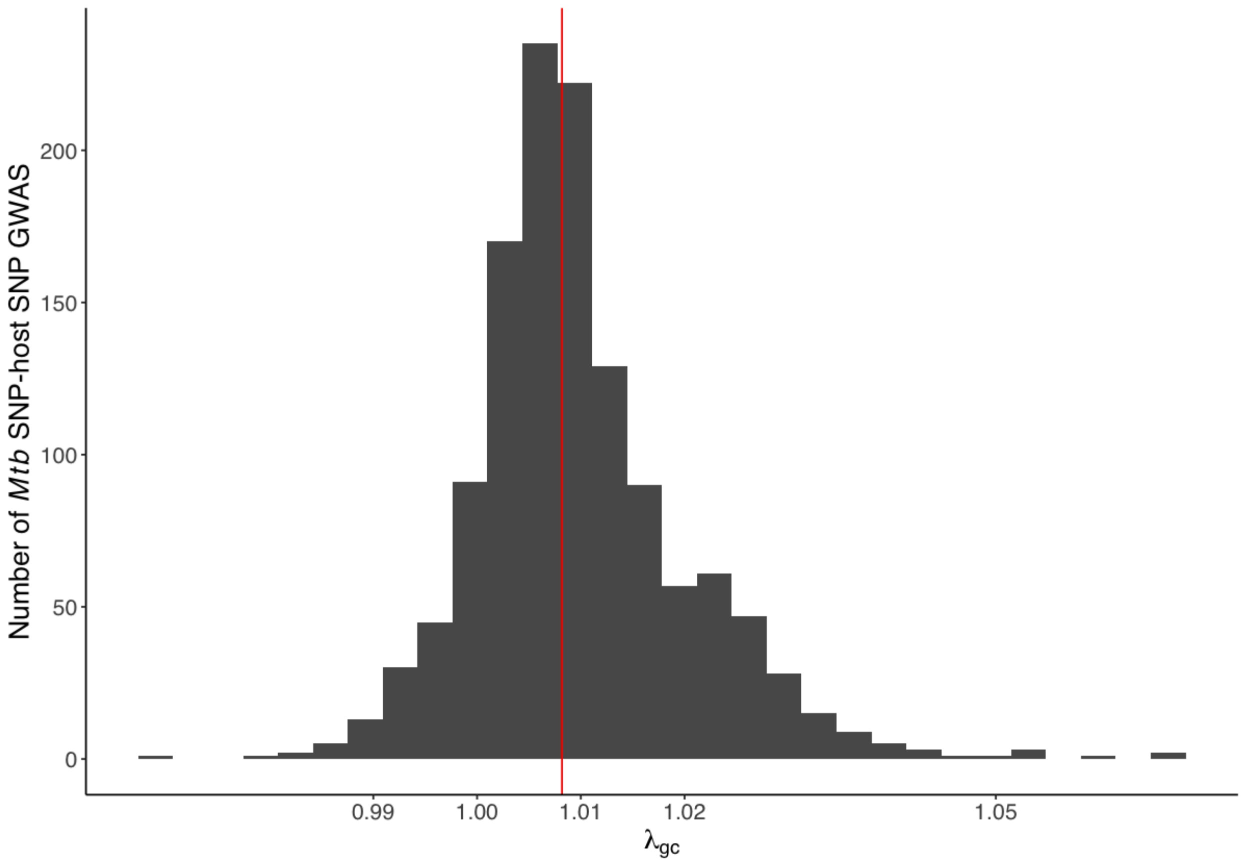

**Figure S2. Empirical estimation of genome-wide significance threshold for the genome-to-genome analysis.** The $-\log_{10}P_{min}$ distribution from 200 permuted genome-to-genome analyses. For each interaction, we randomly assign the presence/absence status of the *Mtb* variant for each individual. The vertical bar in the panel represents the top five percentile of $-\log_{10}P_{min}$ (that is, the estimated empirical genome-wide significance $-\log_{10}P_{sig}$). The dotted gray and black vertical bar represents the common genome-wide significance threshold of $5 \times 10^{-8}$ and the Bonferroni corrected significance threshold after multiple testing ($5 \times 10^{-8}$ /1,267 = $3.95 \times 10^{-11}$).

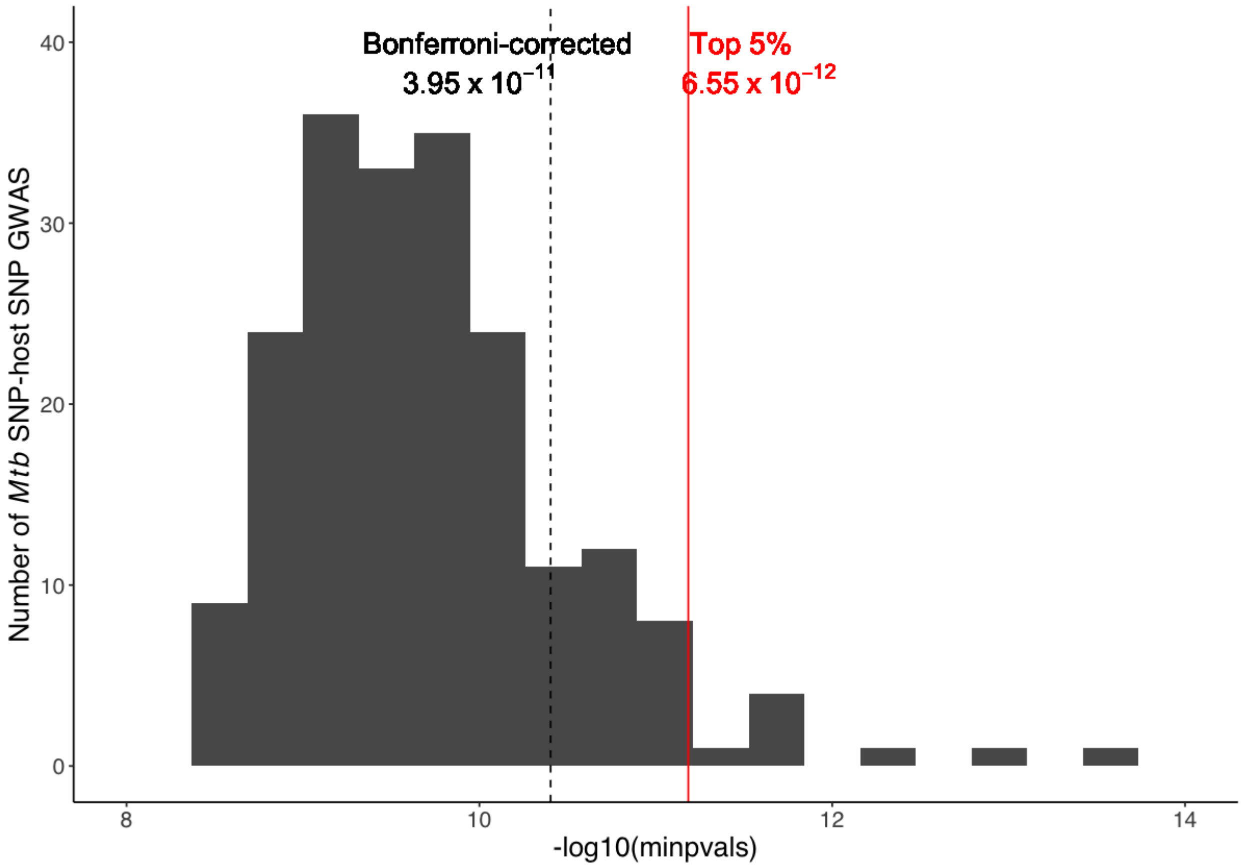

**Figure S3. eQTL effect of rs3130660 for *FLOT1*.** Violin plot of *FLOT1* expression level in (**A**) adipose visceral tissue and (**B**) lung tissue included in the GTEx project, where rs3130660-A is associated with increased *FLOT1* expression. Y-axis shows the normalized gene expression between samples. Boxplots show median (horizontal white bar), 25th and 75th percentiles (lower and upper bounds of the box, respectively). P-value is obtained using FastQTL. Number of individuals included in the analysis for each genotype are in brackets. The figures are modified based on raw data downloaded from the GTEx portal ([https://gtexportal.org/](https://gtexportal.org/)).

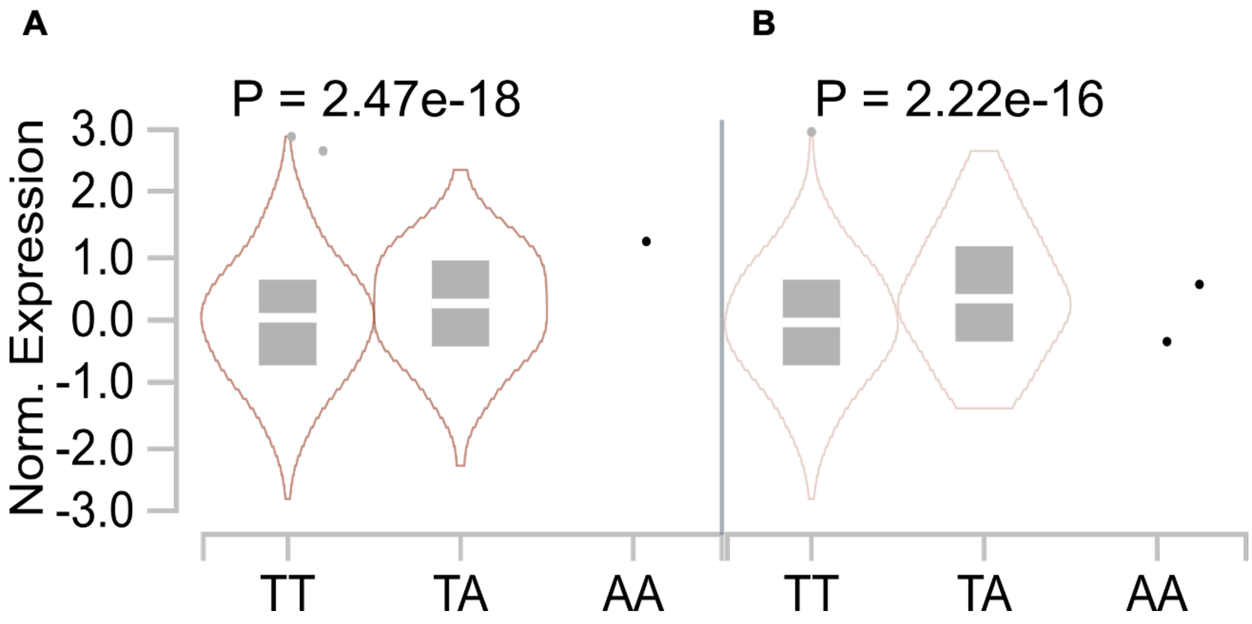

**Figure S4. Correlation between g2g and eQTL p-values in the human *FLOT1* locus.** A correlation plot between -$\log_{10}$(p-value) of the g2g and *FLOT1* eQTL (in lung, GTEx) analyses. The correlation coefficient (Pearson r = 0.88) and the p-value of correlation (P = 1 × $10^{-3917}$) is obtained from a linear regression weighted by the effect size of both g2g and eQTL study. The LD $r^2$ is obtained using the enrolled Peruvian dataset.

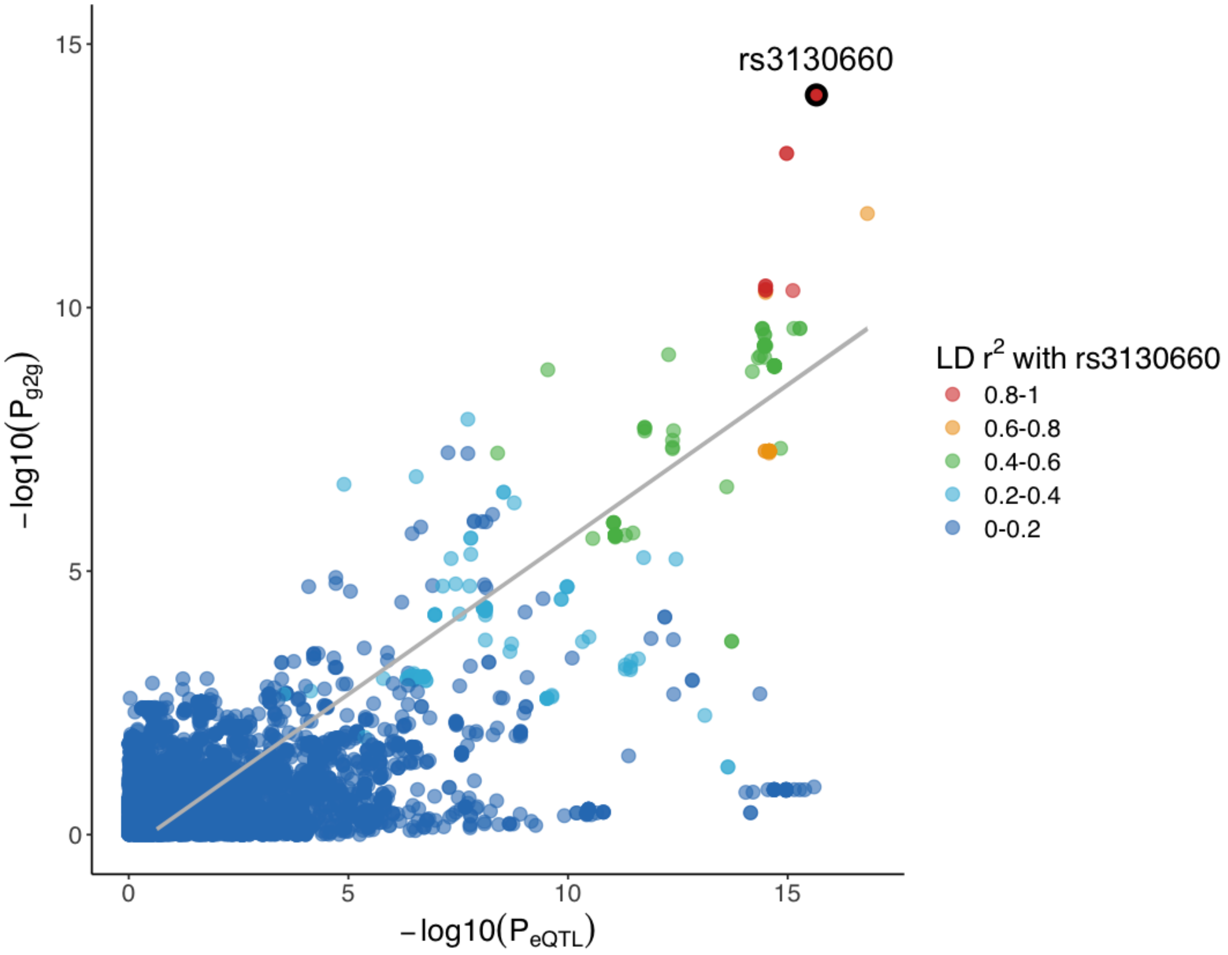

**Figure S5. Colocalization analysis of the host genomic locus that is significantly associated with the *Mtb* genome.** Each grid represents the posterior probability using *coloc*. The color gradient represents from low (posterior probability = 0, gray) to high (posterior probability = 1, red) probability of shared causal variants between the host g2g associations and eQTL among 69 distinct cell types and tissues (108 bulk RNA sequencing datasets) and 49 genes in a ±700kb window of the most significant g2g SNP (rs3130660). The eQTL summary statistics are downloaded from eQTL catalog release 4. Source data are provided in **table S3**.

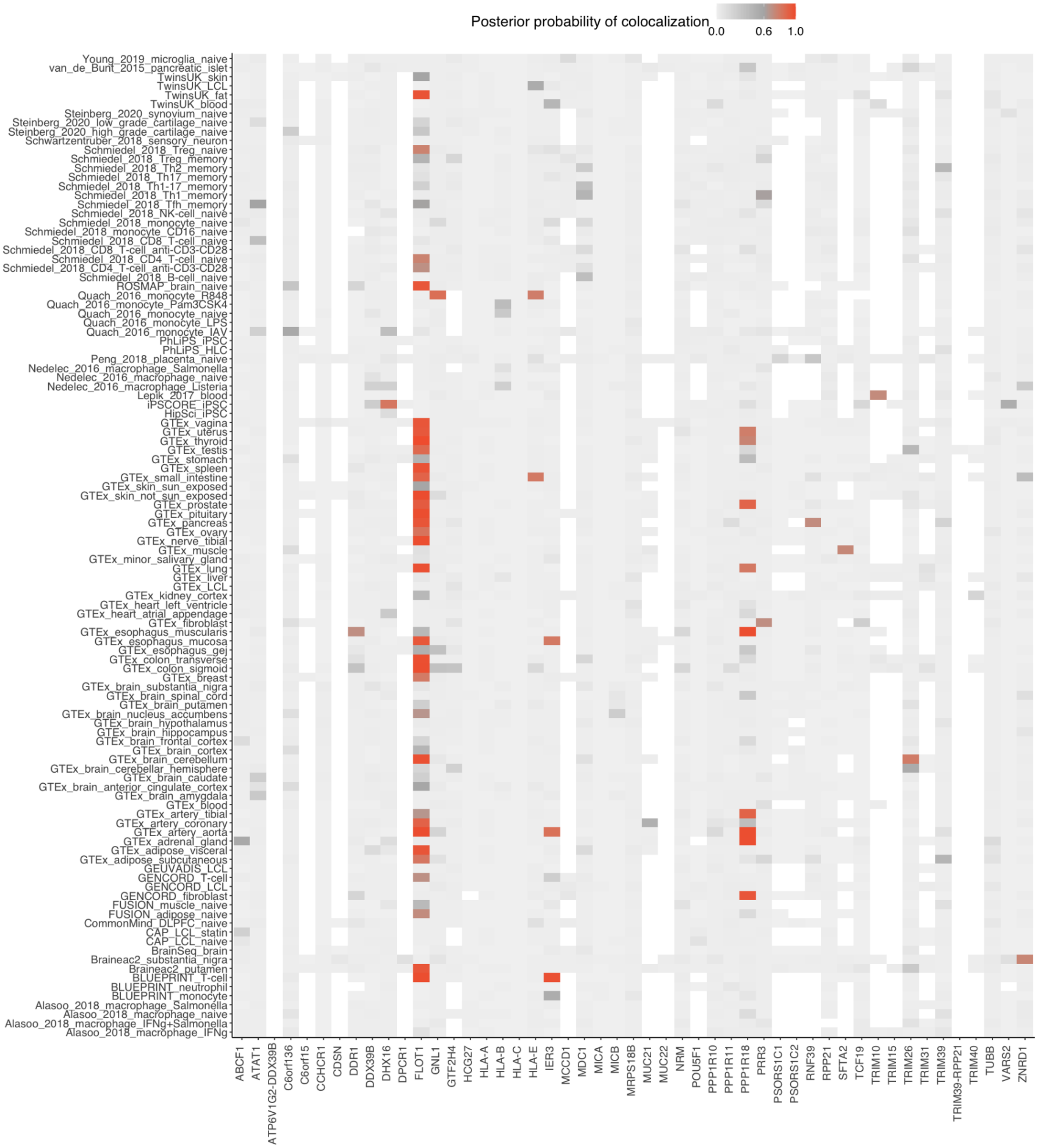

**Figure S6. Odds ratios for association of host variant rs3130660 with the *Mtb* variant (Position 271640) in 1,556 TB patients.** Plot shows parameter estimates (odds ratios, points) and 95% confidence interval (horizontal line segment) for the association of host and *Mtb* genotype at the most significant association site. We run a mixed effect logistic regression where we assume an additive model and correct for various covariates (rows). GRM represents a genetic relationship matrix to account for cryptic relatedness and population structure. First 10 principal components of the *Mtb* genome were constructed using all 2,298 common *Mtb* variants. We constructed 10 alternative *Mtb* principal components of the *Mtb* genome by using all common *Mtb* variants, but excluding the 79 g2g-L2 *Mtb* variants that are significantly associated with the human genome.

| **Covariates** | **OR** | **P–value** |
|---|---|---|
| GRM, age, sex | 1.386 | 9.236e–15 |
| GRM, age, sex, year of sample collection | 1.387 | 8.35e–15 |
| GRM, age, sex,<br>Top 10 Mtb PCs | 1.175 | 3.419e–09 |
| GRM, age, sex,<br>Top 10 Mtb PCs excluding g2g–L2 variants | 1.204 | 7.757e–10 |

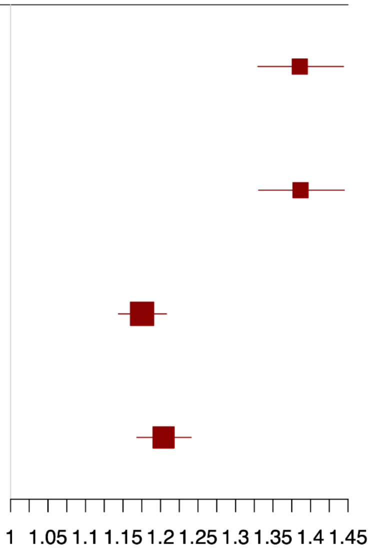

**Figure S7. The constitution of different sublineages for each of the four year of collection (2009 - 2012).** Each color represents different inferred lineages. g2g-L2 (red) represents the unique clade of lineage 2 (other non-g2g L2 sublineages in dark red) identified in this study.

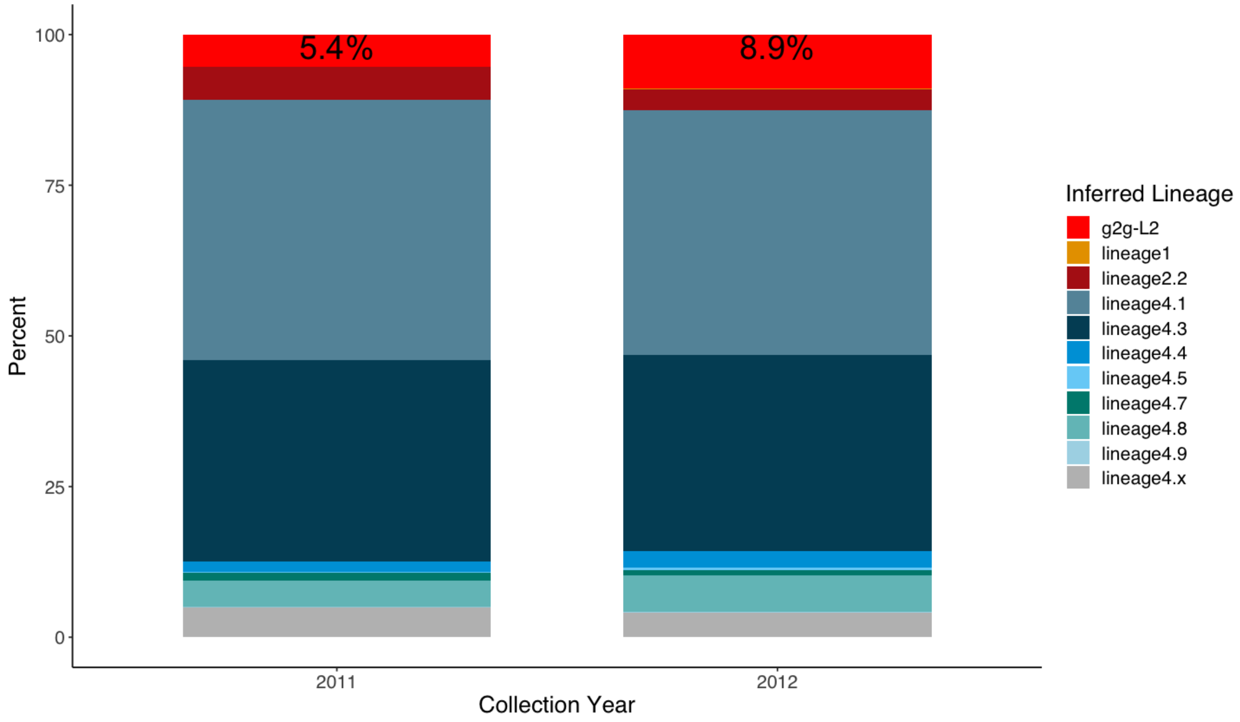

**Figure S8. Geographic distribution of enrolled *Mtb* isolates.** Each point on the map represents the clinic where the *Mtb* sample is collected, and is colored by its inferred lineage. g2g-L2 lineages are defined by the phylogenetic marker (Position 271640). This map shows collected Mtb sublineages were randomly distributed geographically.

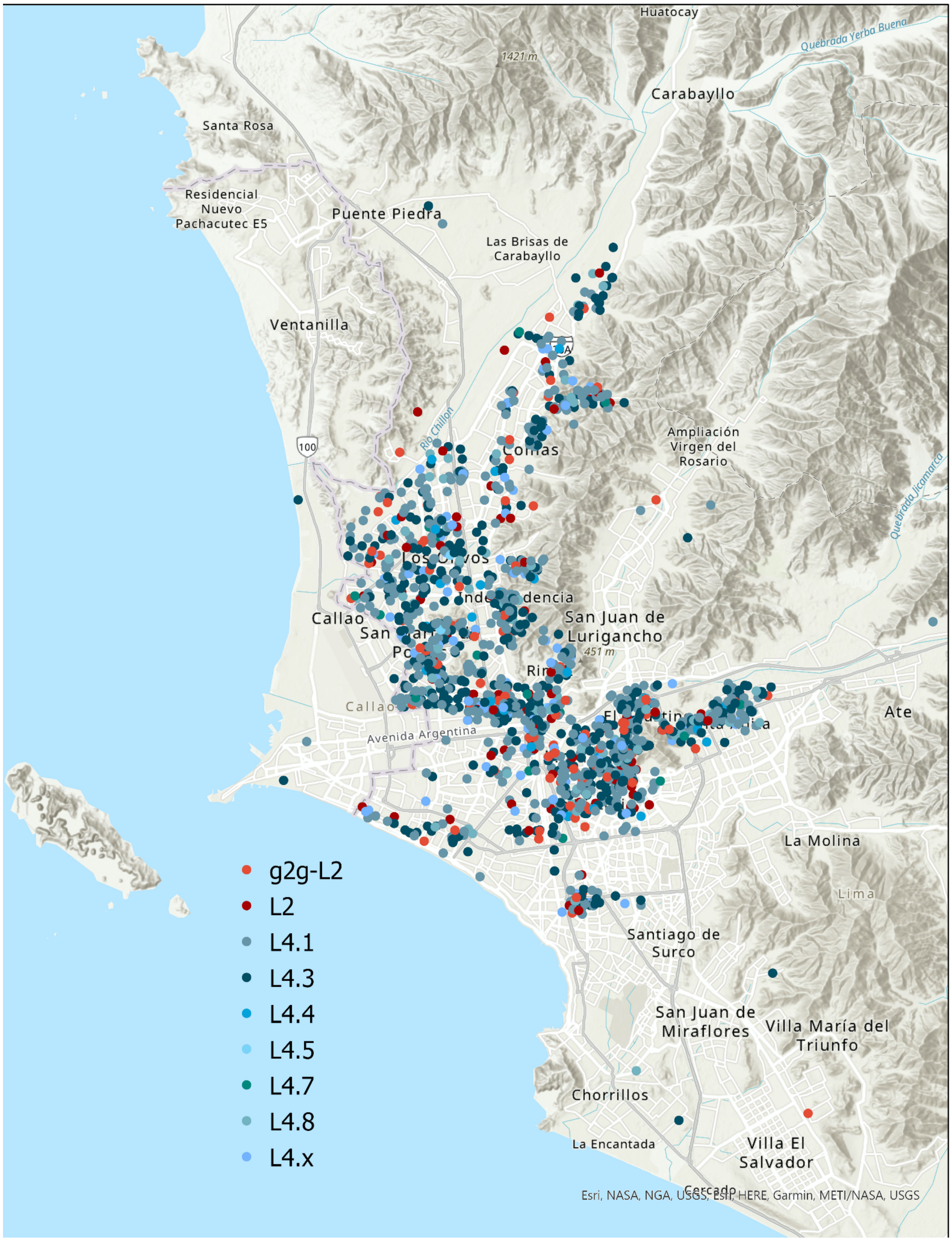

**Figure S9. Genetic ancestry analysis of the 1,556 Peruvian human host genomes.** (**A**) First and second principal components of the Peruvian genomes merged with samples included in the 1000 Genomes Project covering five global populations. (**B**) ADMIXTURE plot of the Peruvian genomes and other global populations (K=5). Each individual is represented as a thin vertical bar. The colors represent the proportion of ancestry assigned to each cluster for each individual. Reference panels are from the 1000 Genomes Project. SAS represents South Asian; EAS represents East Asian; AFR represents African; EUR represents European; LIMA represents Peruvians collected from this cohort, from Lima, Peru.

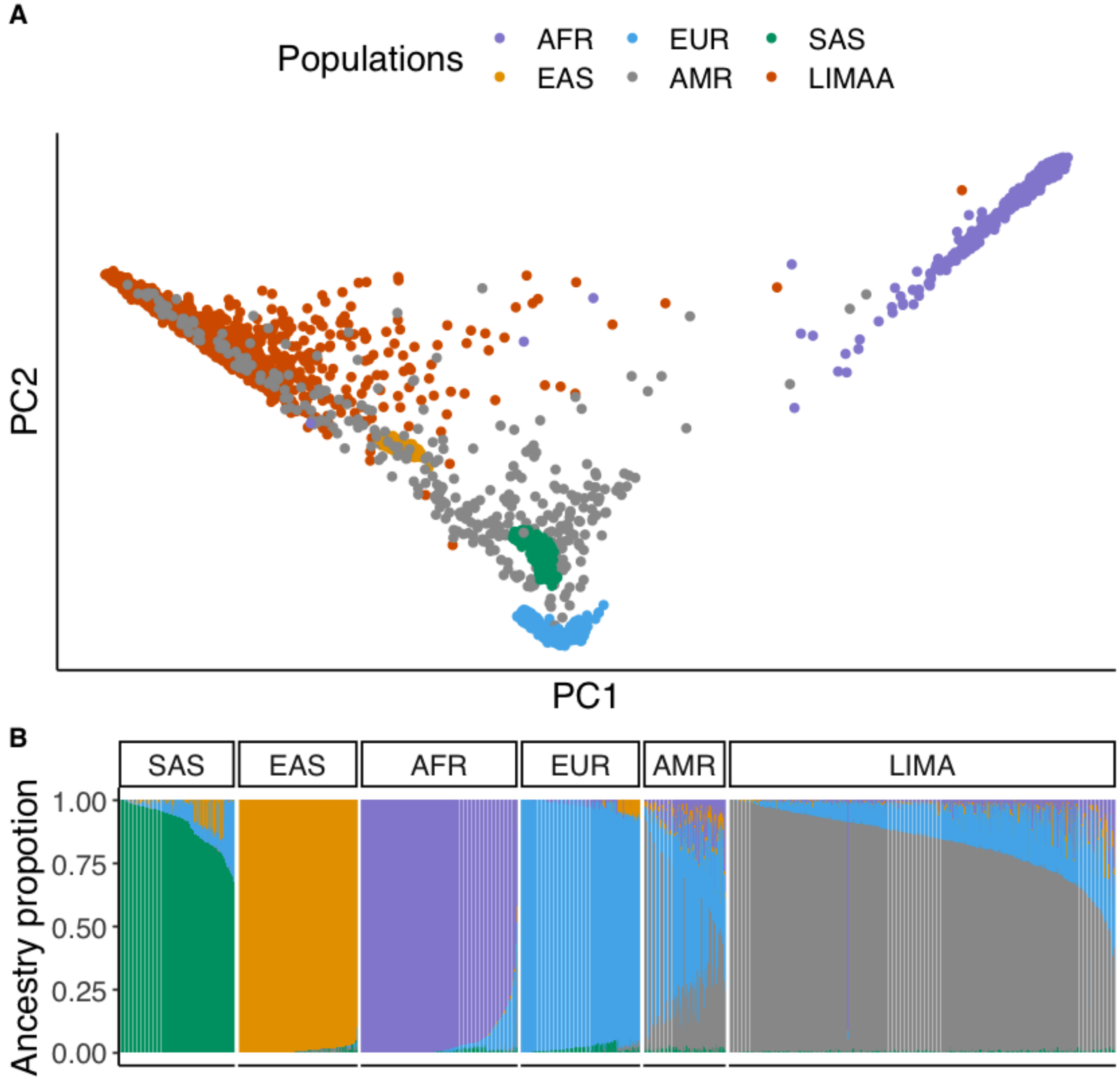

**Figure S10. Genetic structure of Peruvian *Mtb* isolates.** (**A**) A maximum likelihood phylogenetic tree of the collected 1,556 Peruvian *Mtb* isolates. The branches of the tree were colored according to the sublineages. (**B**) Pie chart shows the proportion of different sublineages collected in the study.

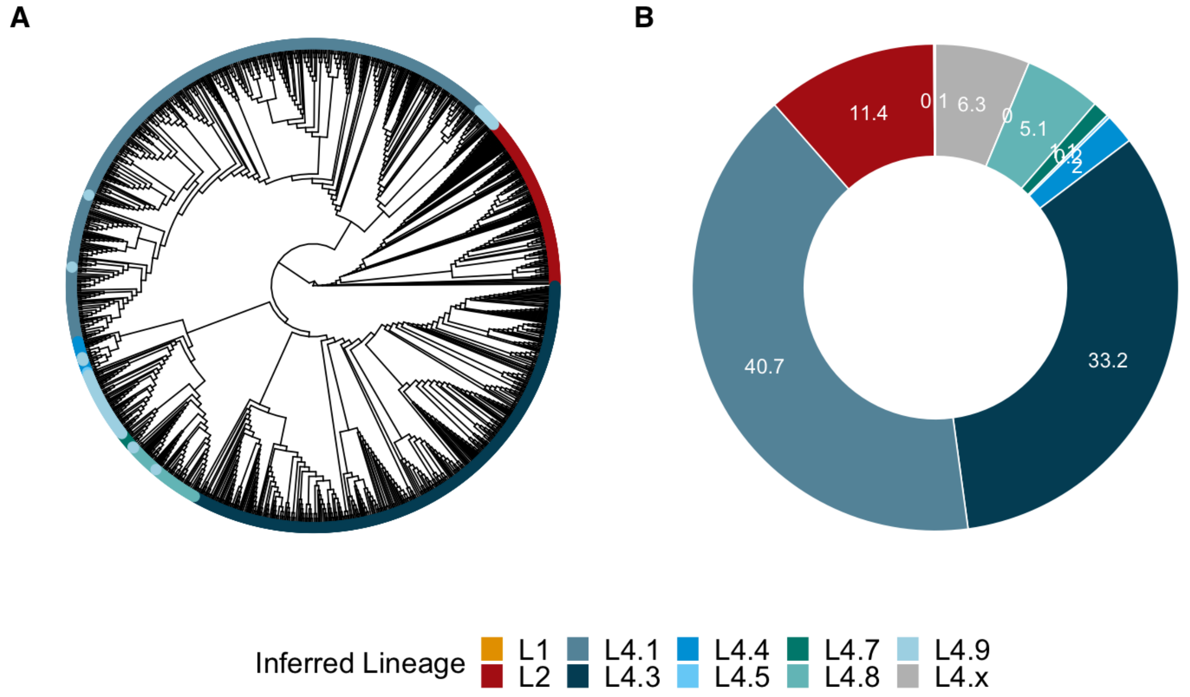

**Figure S11. Human variant-to-*Mtb* lineage genome-wide association study in 1,556 tuberculosis patients.** We tested genomic interplay between *Mtb* lineage and host variants. We performed mixed effects logistic regression assuming an additive model and correcting for host cryptic relatedness, population structure, age and sex. (**A**) Manhattan plot of the GWAS analysis when treating two main *Mtb* lineages as outcome variable (i.e., a binary trait with L2 = 1 and L4=0) prior to imputation. The x-axis denotes the human genomic positions with alternating colors for each chromosome (1-22, X). The y-axis denotes the $-\log_{10}$(P) value from the logistic regression. The red dotted line denotes the genome-wide significance threshold ($5 \times 10^{-8}$). (**B**) Q-Q plot of the GWAS p-values shown in (**A**). (**C**) Regional plot of the MHC region after imputing using a multi-ancestry MHC reference panel. The most significant association is at an intron variant rs3130660 of *FLOT1*. Variants lying within the six classical HLA genes are highlighted in red (HLA-*A*), blue (HLA-*B*), green (HLA-*C*), purple (HLA-*DQA1*), orange (HLA-*DQB1*) and yellow (HLA-*DRB1*). All variants that are not in the classical HLA genes are in gray.

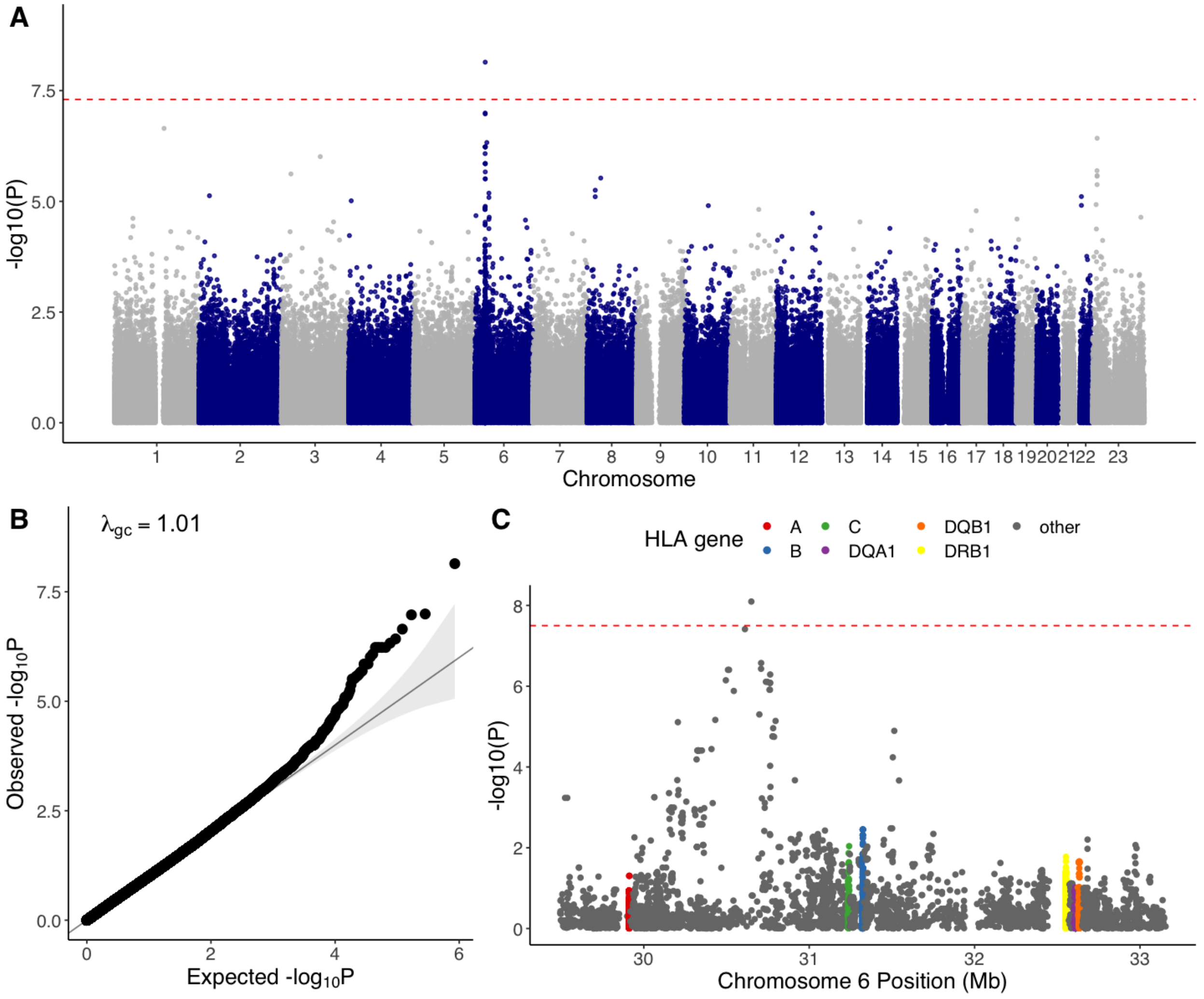

**Figure S12. Peruvian L2 isolates and their global neighbors.** A maximum likelihood phylogenetic tree of Peruvian L2 and 1,000 L2 isolates obtained globally (shown in gray). A total of three Peruvian L2 clades (g2g-L2 in red, clade-B in blue and clade-C in green) are marked. Other Peruvian L2 isolates that do not belong to the three marked clades are shown in pale orange.

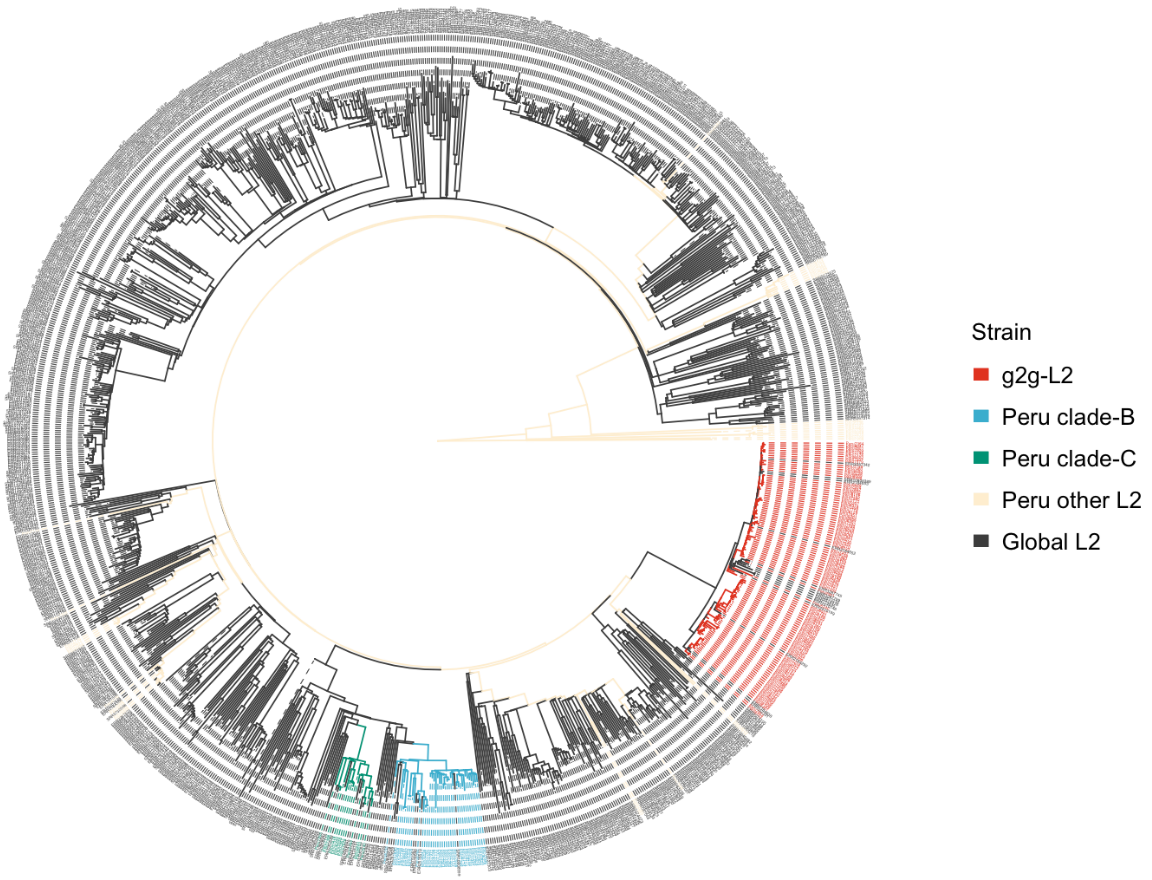

**Figure S13. Geographic distribution of Peruvain L2 *Mtb* isolates.** Each point on the map represents the clinic where the *Mtb* isolate was collected, and is colored by its inferred lineage. g2g-L2 lineages are defined by the phylogenetic marker (Position 271640).

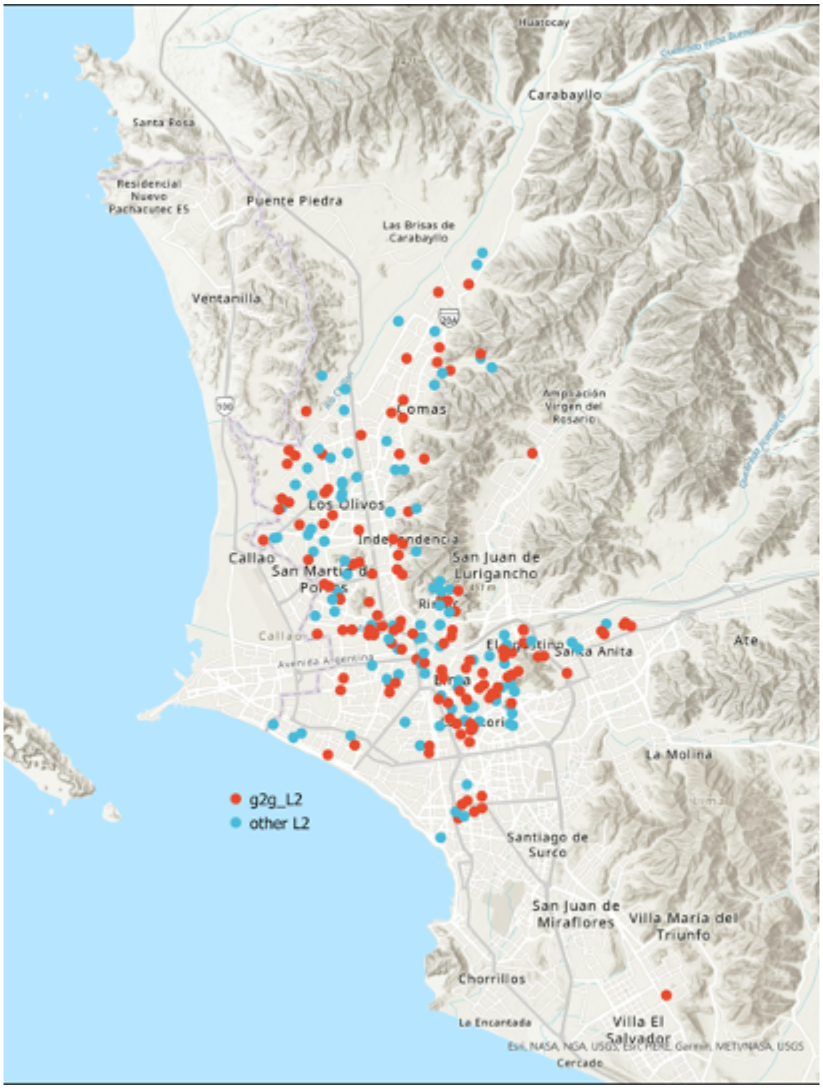

**Figure S14. High degree of correlation between biological replicates assayed via microscopy platform.** Independent biological replicates were assessed through the microscopy platform for each g2g-L2 and non g2g-L2 strain. For each of the seven features [cell length, median cell width, cell area, Nile Red (TRITC), NADA incorporation (FITC), CFP ($F_{420}$-ox) and DNA staining (DAPI)], the Pearson correlation between replicates of each strain are shown. Each point represents a different *Mtb* strain.

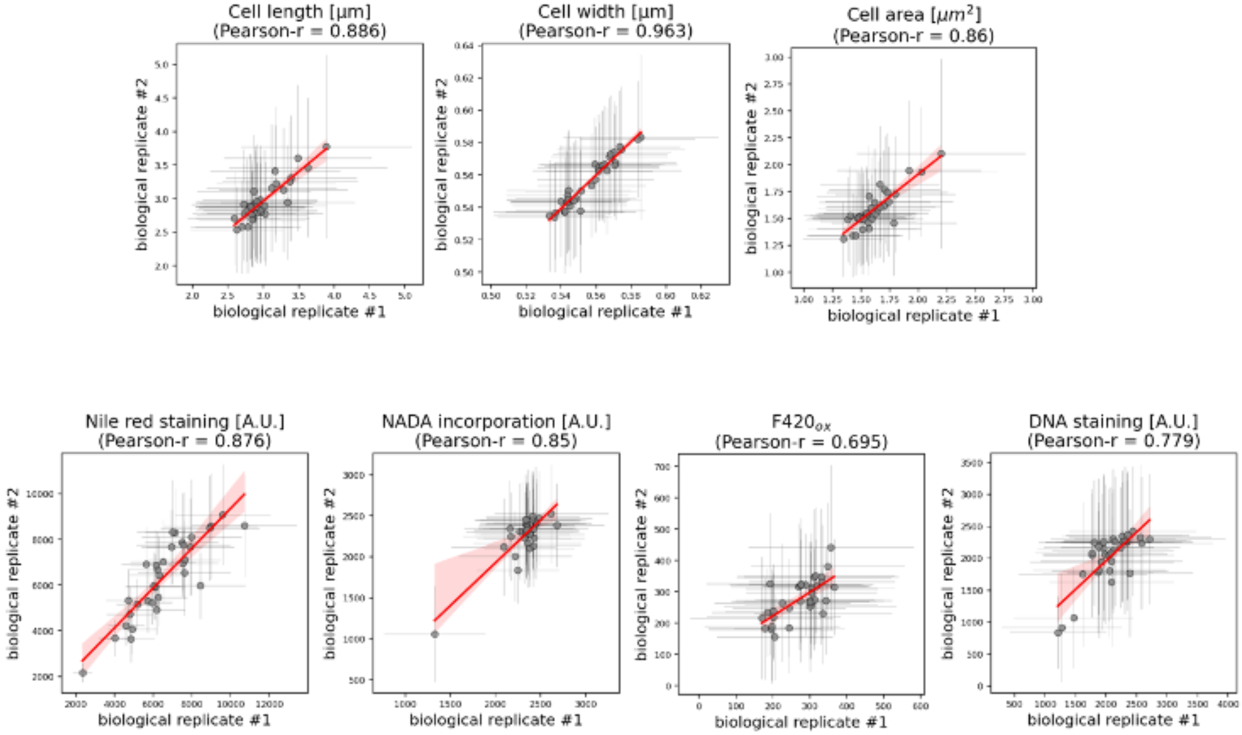

**Figure S15. Functional annotation clustering of *Mtb* genes with nonsynonymous mutations defining g2g-L2 subclade.** Two enriched functional annotation clusters generated by DAVID (https://david-d.ncifcrf.gov/) using 48 *Mtb* genes with nonsynonymous changes in g2g-L2 defining variants (**table S2**). (**A**) shows g2g-L2 genes overlap with ATP binding pathways; and (**B**) shows g2g-L2 genes overlap with redox-related pathways.

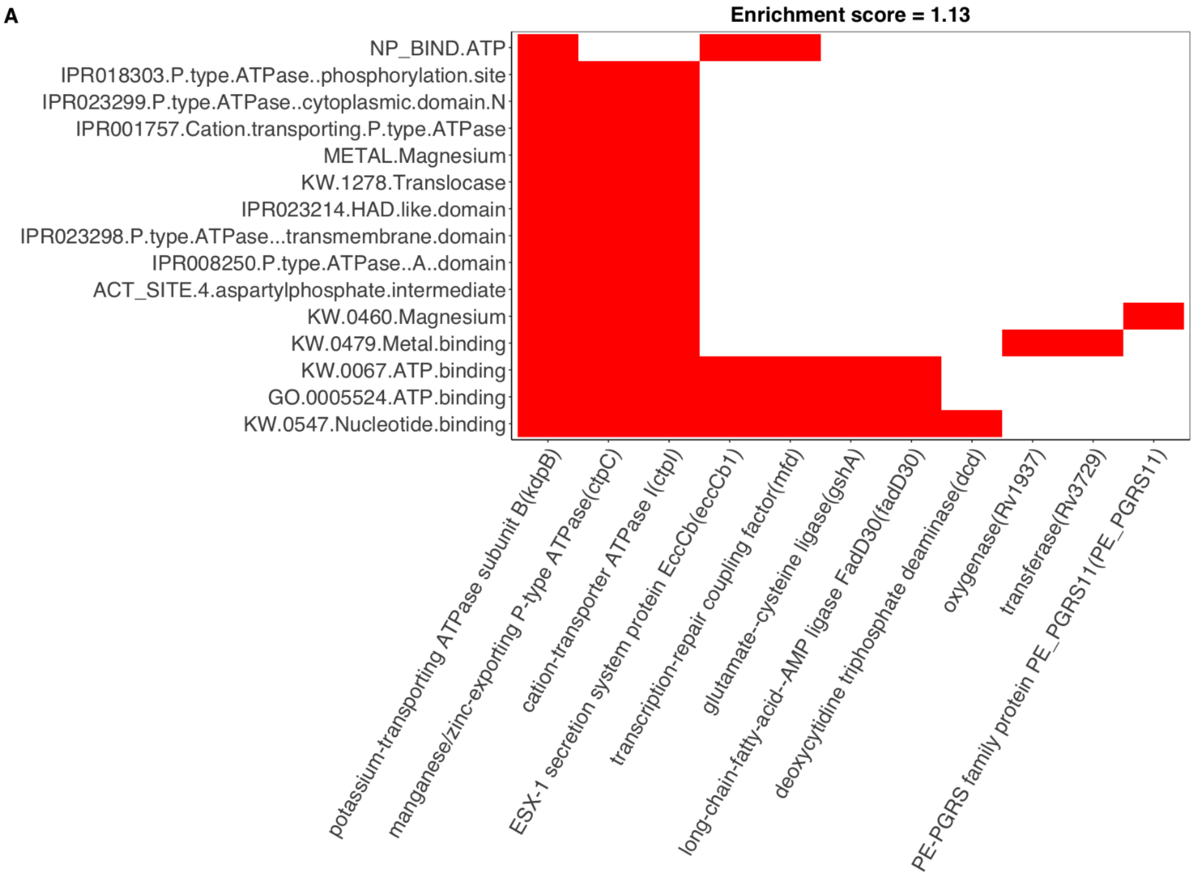


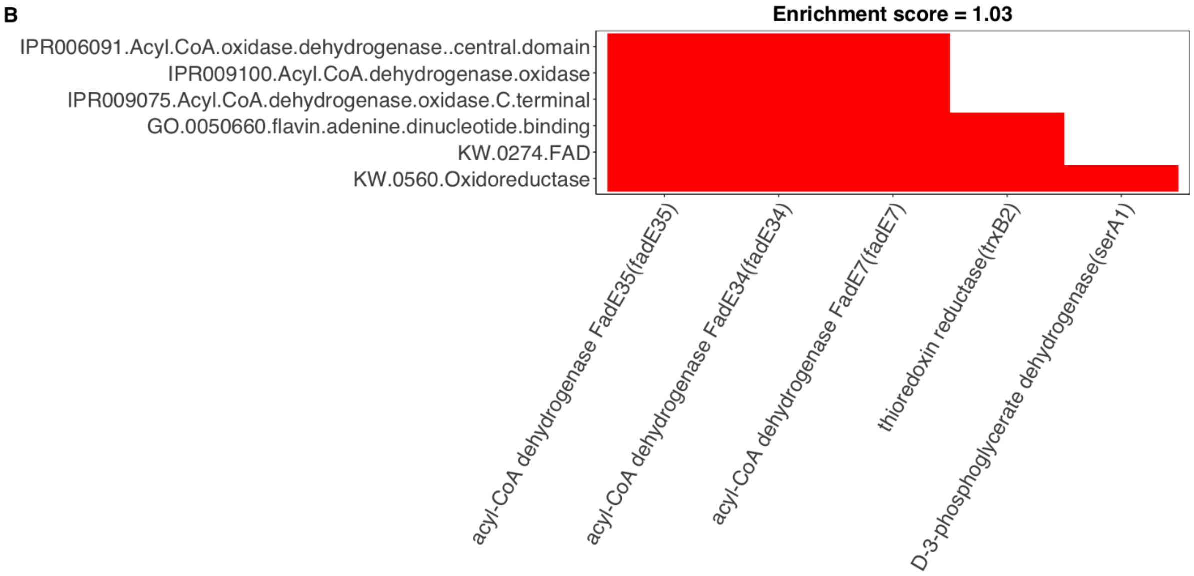
